# Accelerometer-Derived Relative Amplitude and Midlife Cognition: A Cross-Sectional Study

**DOI:** 10.64898/2026.09.15.26361208

**Authors:** Minjee Kim, Morgan Bonham, Fangyu Yeh, Marisol Arredondo, Lauren Opsasnick, Laura Curtis, Stacy C. Bailey, Julia Y. Benavente, Emily H. Ho, Paul Chung, Danielle Wallace, Mary Kwasny, Jeffrey A. Linder, Michael S. Wolf, Phyllis C. Zee

## Abstract

**Importance:** Lower relative amplitude of the rest-activity pattern has been associated with cognitive decline and incident dementia in older adults, but its relationship with domain-specific cognitive performance during midlife is less well characterized.

**Objective:** To examine cross-sectional associations between accelerometer-derived relative amplitude and fluid and crystallized cognitive performance among midlife adults.

**Design, Setting, and Participants:** Cross-sectional analysis of 851 adults aged 35-64 years enrolled in the MidCog Study and recruited from 13 primary care clinics, including academic practices and federally qualified health centers. Participants completed a 14-day wrist accelerometer recording and the NIH Toolbox Cognition Battery.

**Exposure:** Accelerometer-derived relative amplitude, calculated from the contrast between activity during the most active consecutive 10-hour period and the least active consecutive 5-hour period.

**Main Outcomes and Measures:** Age-and education-adjusted NIH Toolbox T-scores (normative mean = 50, SD = 10) for the Fluid Cognition Composite; attention, executive function, episodic memory, working memory, and processing speed; the Crystallized Cognition Composite; receptive language and expressive language. Multivariable linear regression models sequentially adjusted for sociodemographic characteristics, other rest-activity rhythm metrics, established dementia risk factors, average activity, time in bed, and sleep efficiency.

**Results:** In the final adjusted model, each 1-SD lower relative amplitude was associated with a 2.27-point lower Fluid Cognition Composite T-score (95% CI, 1.15–3.40). Lower relative amplitude was also associated with lower attention by 2.74 points (95% CI, 1.49–4.00), executive function by 1.49 points (95% CI, 0.51–2.47), episodic memory by 2.03 points (95% CI, 0.81–3.25), and processing speed by 1.56 points (95% CI, 0.49–2.64). Relative amplitude was not associated with working memory or crystallized cognitive outcomes.

**Conclusions and Relevance:** Among midlife adults recruited from primary care practices, lower relative amplitude was associated with modestly poorer fluid cognitive performance after adjustment for established dementia risk factors and sleep characteristics. Relative amplitude may represent a scalable behavioral correlate of cognitive variability within the normative range. Longitudinal studies beginning in midlife are needed to establish temporality, predictive value, or modifiability.

**Key Points:** *Question:* Among midlife adults, is lower accelerometer-derived relative amplitude associated with poorer cognitive performance?

*Findings:* In this cross-sectional study of 851 adults aged 35-64 years recruited from primary care practices, each 1-SD lower relative amplitude was associated with a 2.27-point (0.23 SD) lower Fluid Cognition Composite T-score. Lower relative amplitude was also associated with poorer attention, executive function, episodic memory, and processing speed, but not working memory or crystallized cognition.

*Meaning:* Relative amplitude may provide a scalable behavioral correlate of variation in fluid cognitive performance during midlife. Longitudinal studies are needed to determine whether it predicts subsequent cognitive decline.

## Introduction

Dementia prevalence and its societal costs are projected to rise substantially over the coming decades.^1^ Prevention efforts increasingly emphasize a life-course approach, with midlife representing a potentially important period for modifying trajectories of cognitive aging.^2–5^ Cognitive aging is not uniform across domains. Fluid abilities, including attention, processing speed, executive function, and working memory, may begin to decline during the 30s and 40s,^6,7^ whereas crystallized abilities such as vocabulary and language are generally preserved until later life.^8^ Scalable behavioral measures associated with variation in fluid cognition could help identify potentially relevant processes before clinically apparent cognitive impairment develops.

Wrist accelerometry provides an objective, low-burden method for characterizing 24-hour rest– activity rhythms (RARs) in free-living conditions. In contrast to parametric RAR metrics, nonparametric RAR metrics do not impose assumptions on data “shape” and instead rely on their statistical properties to capture complementary features of daily activity patterns. These complementary metrics include amplitude, day-to-day stability, and within-day fragmentation. Relative amplitude quantifies the contrast between activity during the most active 10-hour period and the least active 5-hour period, with lower values indicating a weaker distinction between active and rest periods. Relative amplitude may be also be a useful indicator of cognitive vulnerability; prior studies in older adults have prospectivelly linked lower relative amplitude with cognitive decline, mild cognitive impairment, and incident dementia.^9–11^

Evidence earlier in adulthood remains limited. A prospective UK Biobank study of adults aged 43–79 years associated lower relative amplitude with subsequent mild cognitive impairment or dementia, but analyses of the full sample and broad age strata (<70 and ≥70 years) did not isolate midlife.^12^ A separate UK Biobank analysis associated lower relative amplitude with slower reaction time but did not use a comprehensive, domain-specific cognitive battery.^13^ Other studies similarly combined middle-aged and older adults or examined selected populations, such as postmenopausal women.^14–17^ Moreover, relative amplitude is associated with several established dementia risk factors and may be influenced by sleep and physical activity.^18–21^ Thus, its independent association with specific cognitive domains during midlife remains unclear.

To address these gaps, we examined cross-sectional associations between accelerometer-derived relative amplitude and domain-specific cognitive performance among adults aged 35–64 years recruited from primary care practices. Relative amplitude was prespecified as the primary exposure because it is the most consistently studied nonparametric RAR metric in relation to cognitive decline and dementia.^9,10,12^ We evaluated associations across fluid and crystallized cognitive domains using sequential models that accounted for sociodemographic characteristics, other RAR metrics, established dementia risk factors, average activity, and accelerometer-derived sleep characteristics. We hypothesized that lower relative amplitude would be associated with lower Fluid Cognition Composite scores and explored associations across individual fluid and crystallized cognitive domains.

## Methods

### Study Design and Participants

The MidCog Study is an ongoing cohort study of English-speaking adults aged 35-64 years recruited from 13 primary care practices across the Greater Chicago area, including academic practices and federally qualified health centers. The study protocol has been published previously.^22^ Patients were ineligible if they had severe and uncorrectable vision, hearing, or cognitive impairment.^23^ Participants completed in-person and telephone interviews with trained research staff, including physical measurements, cognitive testing, and questionnaires assessing health, sleep, mood, and quality of life.

The Northwestern University Feinberg School of Medicine Institutional Review Board approved the study (STU00214736, STU00221343), and all participants provided written informed consent. This cross-sectional analysis included data collected and processed from March 18, 2022, through April 20, 2026. Participants were eligible for the analytic sample if they had valid accelerometer data and at least one NIH Toolbox cognitive outcome.

### Accelerometer Data Collection and Quality Control

Participants wore a water-resistant accelerometer (ActTrust2; Condor Instruments, Brazil) on the nondominant wrist continuously for 14 days. They were instructed to remove the device only for water submersion and to record bedtimes, wake times, and device removal using an event marker and sleep diary. The protocol prespecified accelerometer data collection in 30-second epochs; however, because of device configuration errors, recordings from n = 103 (12.1%) participants were collected in 60-second epochs.

Recording days were defined from noon to noon. We excluded days with more than 4 hours of nonwear. For recordings spanning a daylight-saving time transition (n = 41), the transition day and subsequent 6 days were excluded. Participants were excluded if they contributed fewer than 5 valid recording days (n = 131) or had erroneous recordings caused by device malfunction (n = 14). After accelerometer quality control, 854 participants remained eligible (**Figure 1**).

**Figure 1.**
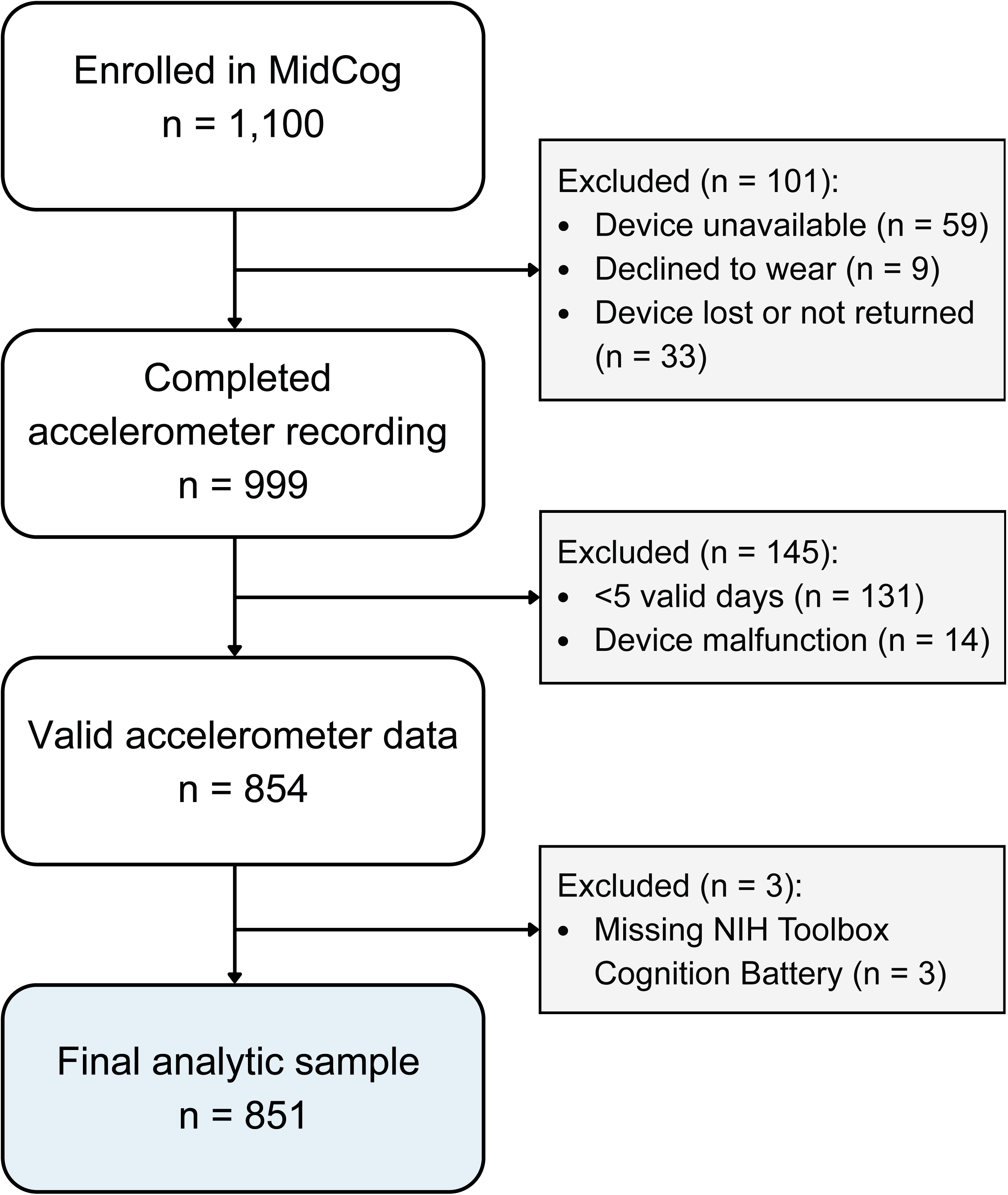
Participant inclusion and exclusion flow diagram for the MidCog Study analytic sample

### Rest–Activity Rhythm Assessment

We calculated nonparametric RAR metrics using the *nparACT* R package (v0.9.1).^24^ Because *nparACT* requires a complete time series, missing activity epochs were handled using 10 stochastic within-person, time-of-day hot-deck completions.^25,26^ The proportion of missing epochs was low (median, 1.3%; IQR, 0.5%-2.5%).

Relative amplitude was prespecified as the primary exposure. It was calculated as the normalized difference between mean activity during the most active consecutive 10-hour period (M10) and the least active consecutive 5-hour period (L5):

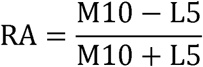

Higher relative amplitude indicates a greater contrast between active and rest periods. Intradaily variability quantifies fragmentation of the activity pattern within a day, with higher values indicating more frequent transitions between activity and rest. Interdaily stability quantifies the similarity of the 24-hour activity pattern across days, with higher values indicating greater day-to-day consistency. Relative amplitude, intradaily variability, and interdaily stability were standardized using common means and SDs across the completed datasets. Participant-level means across datasets were used for descriptive summaries and correlations.

### Sleep Assessment

A trained rater scored each valid accelerometer recording using a standardized hierarchical approach to define rest intervals based on event markers, sleep diaries, light, and activity data.^27^ Each scored recording was independently reviewed by a second trained rater, and disagreements were resolved by consensus during group sessions led by the study principal investigator (M.K.).

After designating rest intervals, ActStudio (Condor Instruments, Brazil) algorithms assigned sleep or wake status to each epoch using 5 immobile minutes to define sleep onset, 0 immobile minutes to define sleep offset, and a wake threshold of 40 activity counts. We defined the longest rest interval in each 24-hour period as the main sleep period. Time in bed, total sleep time, and sleep efficiency ([total sleep time] / [time in bed] × 100) were averaged across all valid main sleep periods. The sleep regularity index quantified the probability of being in the same sleep or wake state at 2 time points 24 hours apart^28^ and was standardized before analysis.

### Cognitive Assessment

We assessed cognitive performance using version 3 of the NIH Toolbox Cognition Battery.^29^ The Fluid Cognition Composite, prespecified as the primary outcome, comprises tests of attention and inhibitory control (Flanker), executive function (Dimensional Change Card Sort), episodic memory (Picture Sequence Memory), working memory (List Sorting Working Memory), and processing speed (Pattern Comparison). The Crystallized Cognition Composite comprises receptive language (Picture Vocabulary) and expressive language (Oral Reading Recognition). Secondary outcomes included the 5 individual fluid domains, the Crystallized Cognition Composite, receptive language, and expressive language. We used the age-and education-adjusted T-scores, standardized to a normative mean of 50 and SD of 10.^29^

### Statistical Analysis

We summarized participant characteristics, accelerometer measures, sleep characteristics, and cognitive scores using medians and interquartile ranges for continuous variables and counts and percentages for categorical variables. To characterize potential selection related to accelerometer data availability, we compared baseline demographic, clinical, and cognitive characteristics between enrolled participants with and without a valid accelerometer recording. Group differences were summarized using absolute standardized mean differences without null-hypothesis significance testing.^30^

We used multivariable linear regression to estimate the difference in cognitive T-score associated with a 1-SD higher relative amplitude. The Fluid Cognition Composite was the primary outcome; individual fluid domains and crystallized outcomes were secondary. Four sequential models were estimated. Model 1 (unadjusted) included relative amplitude as the sole predictor.

Model 2 additionally included intradaily variability, interdaily stability, age, sex, education, and recruitment site type as covariates. Model 3 additionally adjusted for established midlife dementia risk factors, including hearing impairment, hypercholesterolemia, depressive symptoms, average daily activity, diabetes, current smoking, hypertension, obesity, and excessive alcohol use.^31^ Model 4 additionally included time in bed and sleep efficiency. **eTable 1** provides definitions of all study variables.

We analyzed each of the 10 completed RAR datasets separately. Regression coefficients, variances, marginal means, and contrasts were combined using Rubin’s rules; R² was pooled across datasets.^32^ Common centering and scaling constants were applied across datasets. For each outcome and model, analyses were restricted to participants with complete outcome and covariate data across all datasets.

Because the sleep regularity index was strongly correlated with relative amplitude (Pearson *r* = 0.74) and captures an overlapping dimension of behavioral rhythmicity, it was not included in the primary models. Model 5 added the sleep regularity index to Model 4 as a sensitivity analysis. We assessed multicollinearity in Models 4 and 5 using variance inflation factors (VIFs) and adjusted generalized VIFs, with VIF values greater than 5 considered potentially problematic.^33^ We examined potential effect modification by age, sex, and recruitment site type by adding one multiplicative interaction term at a time to Model 4. As an exploratory analysis, relative amplitude was modeled in quartiles to describe the exposure–response pattern without assuming linearity. Quartile cut points were defined using the pooled relative-amplitude distribution and applied consistently across the imputed datasets. These analyses were considered secondary and hypothesis-generating.

A 2-sided *P*<.05 was considered statistically significant for the prespecified Fluid Cognition Composite analysis. We applied Benjamini-Hochberg correction across the 8 secondary outcomes separately within each fitted model, separately for each interaction moderator, and separately for the quartile trend tests. Analyses were conducted using R (v4.6.1).^34^

## Results

### Sample Description

Of 1,100 participants enrolled in the MidCog study, 854 had valid accelerometer data and 246 did not (**Figure 1**). Across measured characteristics, absolute standardized mean differences ranged from 0.02 to 0.33. The largest differences involved recruitment site type (0.19) and cognitive scores (range, 0.14-0.33); participants without valid accelerometer data were more frequently recruited from federally qualified health centers and had lower cognitive scores than those with valid data (**eTable 2**). Three participants with valid accelerometer data lacked all NIH Toolbox outcomes and were excluded, yielding a final analytic sample of 851 participants.

Among 851 participants, 338 (39.7%) self-identifed as non-Hispanic Black, 329 (38.7%) as non-Hispanic White, and 114 (13.4%) as Hispanic or Latino. The median age was 53.8 (IQR, 46.0-59.9) years, 62.6% were female, and 23.0% were recruited from federally qualified health centers. The median Fluid Cognition Composite T-score was 48 (IQR, 42-55).

Participants contributed a median of 14 (IQR, 12-15) valid days of accelerometer recording. Median relative amplitude was 0.9 (IQR, 0.8-0.9). Median time in bed was 7.6 (IQR, 7.0-8.3) hours, total sleep time was 6.9 (IQR, 6.2-7.5) hours, sleep efficiency was 90.4 (IQR, 87.4-92.9)%, and sleep regularity index was 79.5 (IQR, 70.6-85.5). Participant characteristics are shown in **Table 1**; correlations among continuous variables are presented in **eFigure 1**.

**Table 1.**
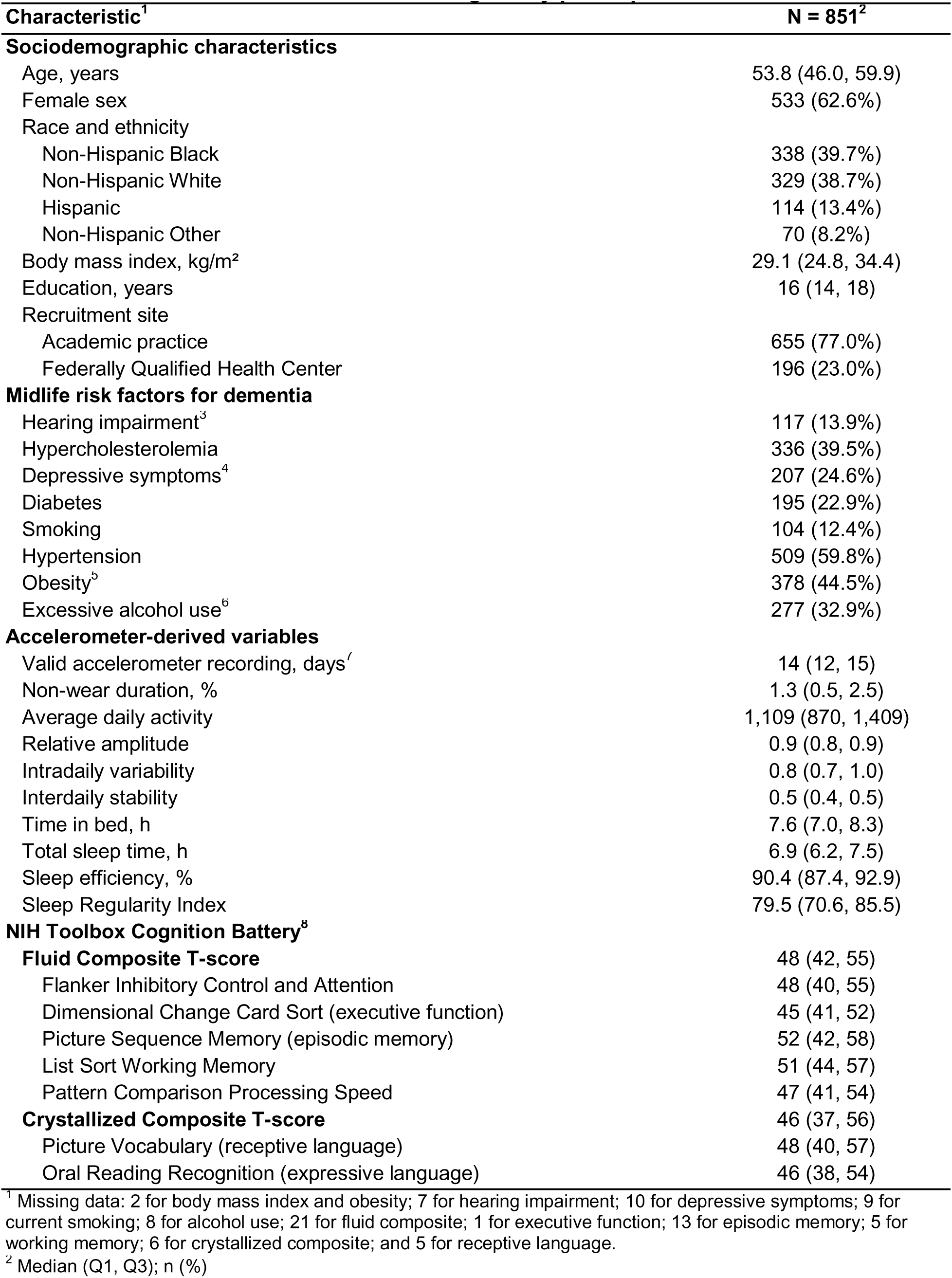
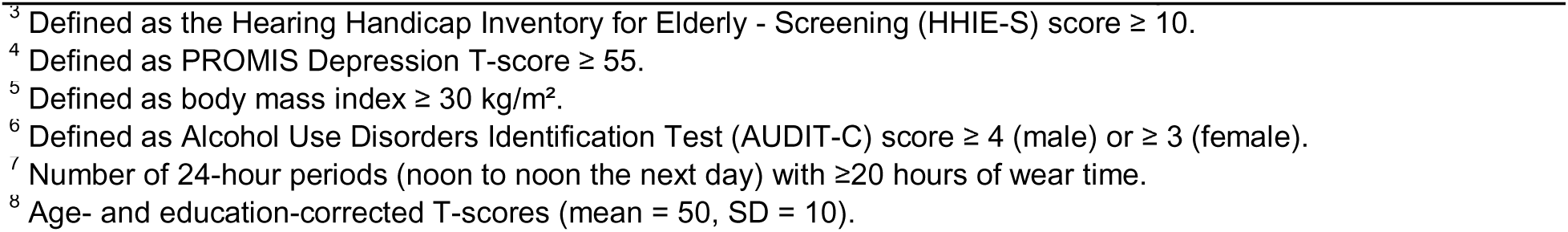
Baseline characteristics of MidCog Study participants.

### Relative Amplitude and Cognitive Performance

In unadjusted analyses (Model 1), lower relative amplitude was associated with lower scores across all cognitive domains (**Figure 2**). After adjustment for sociodemographic characteristics and other RAR metrics in Model 2, lower relative amplitude remained associated with lower Fluid Cognition Composite, attention, executive function, and episodic memory T-scores. Further adjustment for established dementia risk factors in Model 3 produced little change in these estimates.

**Figure 2.**
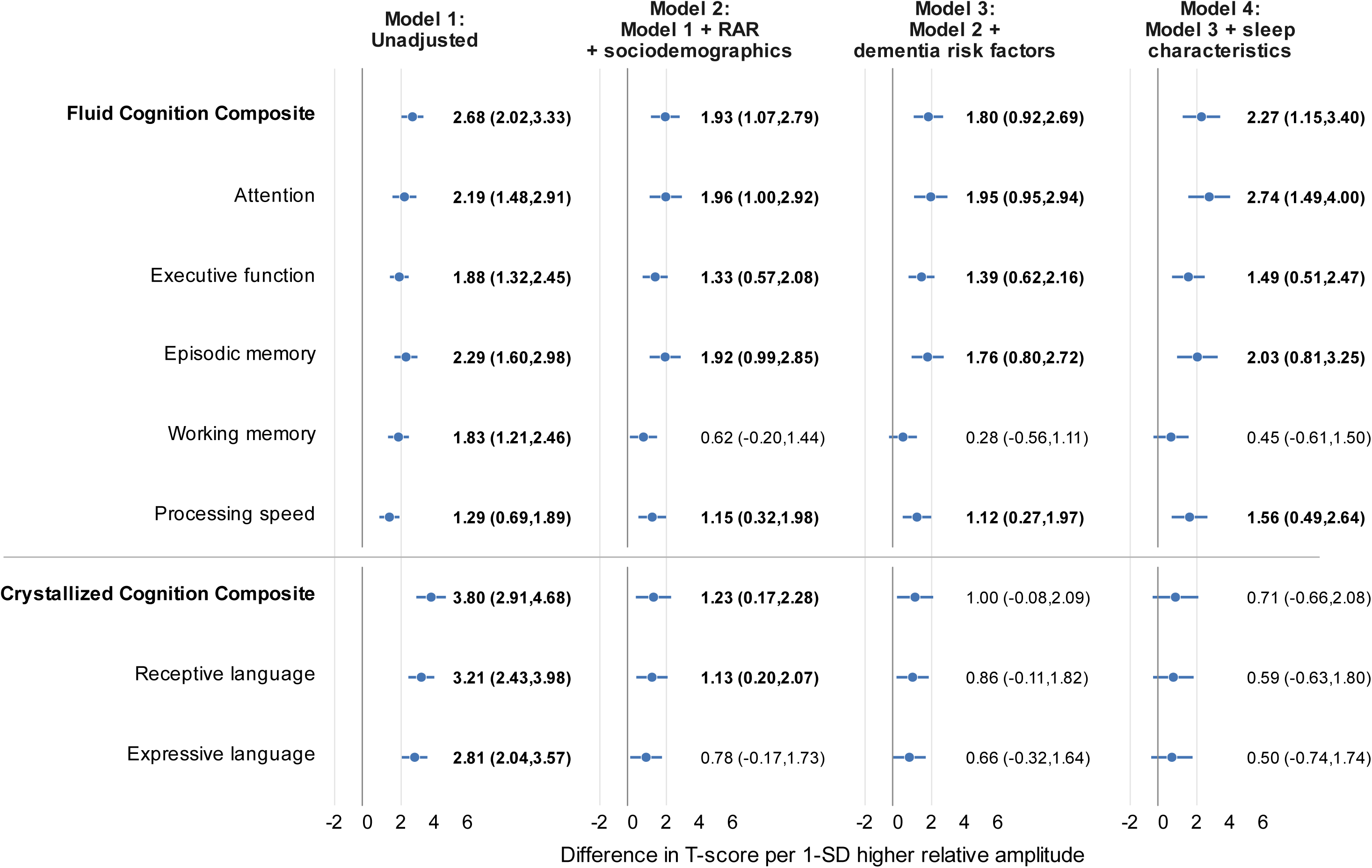
Associations between relative amplitude and NIH Toolbox cognitive domain T-scores across sequentially adjusted models Point estimates and 95% CIs show differences in age-and education-corrected T-scores per 1-SD higher relative amplitude. Numeric labels show coefficients and 95% CIs. Bold labels indicate statistical significance under the prespecified testing procedure: *P* < .05 for the Fluid Cognition Composite and Benjamini–Hochberg false-discovery-rate-adjusted *P* < .05 for the 8 secondary outcomes within each model. RAR indicates rest–activity rhythm. Model 1 was unadjusted. Model 2 additionally included interdaily stability, intradaily variability, age, sex, education, and recruitment site type. Model 3 additionally included hearing impairment, hypercholesterolemia, depressive symptoms, average daily activity, diabetes, current smoking, hypertension, obesity, and excessive alcohol use. Model 4 additionally included time in bed and sleep efficiency.

In Model 4, which additionally adjusted for time in bed and sleep efficiency, each 1-SD lower relative amplitude was associated with an estimated mean of 2.27-point lower Fluid Cognition Composite T-score (95% CI, 1.15–3.40). Among the secondary fluid outcomes, each 1-SD lower relative amplitude was associated with lower attention (β, 2.74; 95% CI, 1.49–4.00), executive function (β, 1.49; 95% CI, 0.51–2.47), episodic memory (β, 2.03; 95% CI, 0.81–3.25), and processing speed (β, 1.56; 95% CI, 0.49–2.64) T-scores. These four secondary associations remained significant after Benjamini-Hochberg correction. Relative amplitude was not associated with working memory, the Crystallized Cognition Composite, receptive language, or expressive language. **Figure 2** shows estimates across the sequential models, and **eTables 3–11** provide complete model results. No evidence of problematic multicollinearity was observed in Model 4 (maximum adjusted generalized VIF = 1.79; **eTable 12**).

### Sensitivity and Exploratory Analyses

Adding the sleep regularity index in Model 5 reduced the Fluid Cognition Composite estimate by 12.7%, from 2.27 to 1.99 T-score points per 1-SD higher relative amplitude. Associations with attention, executive function, and episodic memory remained significant after Benjamini-Hochberg correction, whereas the processing-speed association did not (**eTables 3–11**). Multicollinearity diagnostics for Model 5 remained below the prespecified threshold (**eTable 13**).

For the prespecified Fluid Cognition Composite outcome, there was no evidence that the relative-amplitude association differed by age (*P* for interaction = 0.83), sex (*P* = 0.09), or recruitment site type (*P* = 0.21). Several interaction tests for secondary outcomes were nominally significant, but none remained significant after Benjamini-Hochberg correction (**eTable 14**).

In exploratory quartile analyses, adjusted Fluid Cognition Composite and episodic memory scores were lower in the lowest relative-amplitude quartile than in the highest quartile, with adjusted differences of 4.0 and 5.1 points, respectively. Patterns across quartiles were otherwise inconsistent and did not provide clear evidence of a threshold association (**eFigure 2**).

## Discussion

In this cross-sectional study of midlife adults recruited from primary care practices, lower accelerometer-derived relative amplitude was associated with lower Fluid Cognition Composite scores and poorer performance in several fluid cognitive domains. Associations with attention, executive function, episodic memory, and processing speed persisted after adjustment for sociodemographic characteristics, other rest–activity rhythm metrics, dementia risk factors, average activity, time in bed, and sleep efficiency. Relative amplitude was not associated with crystallized cognition in the primary continuous model.

These findings add to evidence linking altered rest–activity rhythms with cognitive health. Prospective studies in older adults have associated lower relative amplitude with cognitive decline, mild cognitive impairment, and incident dementia.^10,21,35,36^ Evidence during midlife is more limited. Among 194 postmenopausal women with a mean age of 59 years, lower relative amplitude was associated with worse processing speed.^17^ In the Baltimore Longitudinal Study of Aging, which included cognitively healthy adults aged 50 years and older, later timing of least active period was associated cross-sectionally with poorer memory, whereas greater interdaily stability predicted slower longitudinal memory decline.^15^ Our findings extend this literature to a primary care-based cohort spanning ages 35-64 and suggest that lower relative amplitude is more consistently associated with fluid rather than crystallized cognitive performance.

The observed associations were modest. Each 1-SD lower relative amplitude corresponded to approximately 1.5-2.7 points lower T-scores across the associated fluid domains. Average cognitive scores nevertheless remained within normative ranges across relative-amplitude quartiles. Thus, relative amplitude appears to capture cross-sectional variation in cognitive performance within this midlife population rather than established cognitive impairment. Longitudinal studies should clarify the clinical significance of these modest differences by examining whether lower relative amplitude in midlife predicts subsequent domain-specific cognitive decline.

Relative amplitude should be interpreted as a measure of the observed rest–activity pattern rather than a direct measure of endogenous circadian amplitude. Because it is calculated from the contrast between activity during the most active 10 hours and least active 5 hours, relative amplitude reflects the combined influence of circadian regulation, sleep, magnitude and timing of physical activity, and social and environmental schedules.^37,38^ In our study, associations persisted after adjustment for average activity, time in bed, and sleep efficiency and in a sensitivity analysis that also included the sleep regularity index. Thus, these measured activity and sleep characteristics did not fully account for the findings, although residual confounding and overlap among these behavioral measures remain possible.

We did not observe independent associations of interdaily stability or intradaily variability with cognitive performance. This differs from findings in some older or clinically enriched cohorts, in which lower interdaily stability and higher intradaily variability have been associated with cognitive decline, dementia risk, or structural brain alterations.^10,39–41^ This heterogeneity suggests that the cognitive relevance of individual RAR metrics may differ by population and stage of cognitive aging.

Several pathways may explain the association between altered RARs and cognitive performance. At a functional level, disrupted or misaligned RARs may reduce circadian support for alertness and cognitive efficiency, particularly in fluid domains such as attention, processing speed, and memory encoding.^42^ Over longer periods, altered RARs may be related to metabolic, inflammatory, vascular, or neurodegenerative processes. Recent imaging studies have linked RAR characteristics to white matter microstructure, cortical thickness, medial temporal volumes, and subsequent brain volume loss.^40,43,44^ Reverse causation also remains possible, because early neuropathology could disrupt neural systems governing sleep, arousal, and circadian organization, resulting in altered or more fragmented activity patterns.

This study has several limitations. Its cross-sectional design precludes conclusions about temporality or causality. Relative amplitude is a behavioral RAR measure and does not isolate endogenous circadian timing or amplitude. Residual confounding by unmeasured social or environmental factors also cannot be excluded. We assessed some dementia risk factors by self-report, and information on traumatic brain injury history, an established midlife dementia risk factor,^31^ was unavailable. Requiring valid accelerometer data may have introduced selection bias. Participants without valid accelerometer data had lower cognitive scores and were more frequently recruited from federally qualified health centers. Because relative amplitude could not be estimated among excluded participants, the direction and magnitude of any resulting selection bias are uncertain, and generalizability to patients with lower cognitive performance or receiving care in safety-net settings may be limited. Finally, analyses across multiple correlated cognitive outcomes increase the possibility of chance findings, although the prespecified secondary models were controlled using the Benjamini-Hochberg procedure.

In conclusion, lower accelerometer-derived relative amplitude was associated with modestly poorer fluid cognitive performance in midlife adults, even after adjustment for measured activity, time in bed, sleep efficiency, and established dementia risk factors. These findings support relative amplitude as a scalable behavioral correlate of cognitive variability. Longitudinal studies beginning in midlife are needed to determine whether lower relative amplitude precedes domain-specific cognitive decline, reflects early changes in brain health, or both; intervention studies are required to establish whether modifying rest-activity patterns improves cognitive trajectories.

## Data Availability

The de-identified individual participant data that underlie the results reported in this article will be made available to qualified researchers upon reasonable request.

## Acknowledgments

We thank the MidCog study participants and research staff. MK and FY had full access to the data and can take responsibility for its integrity and accuracy in the analysis. MK affirms that the manuscript is an honest, accurate, and transparent account of the study being reported, and no important aspects of the study have been omitted. Artificial intelligence (AI) tools (Codex GPT-5.6 Sol, OpenAI) were used to revise R codes for multiple imputation and figure formatting and to review the language for readability and flow. All AI-assisted content was reviewed, edited, and verified by the authors.

