## Supplement for "Accelerometer-Derived Relative Amplitude and Midlife Cognition: A Cross-Sectional Study"

**eTable 1.** Study variables and operational definitions

**eTable 2.** Characteristics of participants with and without valid accelerometer data

**eFigure 1.** Correlations among rest–activity rhythm, activity, sleep, and demographic measures

**eTable 3.** Associations between relative amplitude and NIH Toolbox Fluid Cognition Composite T-scores

**eTable 4.** Associations between relative amplitude and NIH Toolbox attention T-scores

**eTable 5.** Associations between relative amplitude and NIH Toolbox executive function T-scores

**eTable 6.** Associations between relative amplitude and NIH Toolbox episodic memory T-scores

**eTable 7.** Associations between relative amplitude and NIH Toolbox working memory T-scores

**eTable 8.** Associations between relative amplitude and NIH Toolbox processing speed T-scores

**eTable 9.** Associations between relative amplitude and NIH Toolbox Crystallized Cognition Composite T-scores

**eTable 10.** Associations between relative amplitude and NIH Toolbox receptive language T-scores

**eTable 11.** Associations between relative amplitude and NIH Toolbox expressive language T-scores

**eTable 12.** Multicollinearity diagnostics for Model 4 across cognitive outcomes

**eTable 13.** Multicollinearity diagnostics for Model 5 across cognitive outcomes

**eTable 14.** Interactions between relative amplitude and age, sex, and recruitment site type

**eFigure 2.** Adjusted marginal mean NIH Toolbox cognitive domain T-scores by relative amplitude quartile

**eTable 1. Study variables and operational definitions**

| **Variable** | **Definition and measurement** |
| --- | --- |
| **Demographics** | |
| Age | Difference between the date of in-person interview and date of birth. |
| Sex | Self-reported sex, coded as female vs non-female for regression models. |
| Race and ethnicity | Self-identified race and ethnicity were assessed via structured questionnaire items. Categories were constructed based on responses and collapsed into four groups due to small cell sizes (<5%): Hispanic, non-Hispanic White, non-Hispanic Black, and Other. |
| Education | Years of education completed. |
| Recruitment site | Primary care recruitment setting: academic internal medicine practice or federally qualified health center. |
| **Rest-activity metrics** | |
| Relative amplitude | Normalized difference between activity during the 10 consecutive most active hours (M10) and the five consecutive least active hours (L5); standardized (z-scored) before regression analysis. |
| M10 | Average activity during the 10 consecutive most active hours of the 24-hour rest-activity rhythm profile. |
| L5 | Average activity during the five consecutive least active hours of the 24-hour rest-activity rhythm profile. |
| Intradaily variability | Measure of within-day fragmentation of the rest-activity rhythm; standardized (z-scored) before regression analysis. |
| Interdaily stability | Measure of day-to-day stability of the 24-hour rest-activity rhythm profile; standardized (z-scored) before regression analysis. |
| Average daily activity | Accelerometer-measured activity counts averaged over all valid 24-hour periods (noon-to-noon) of recording; standardized (z-scored) before analysis. |
| **Sleep characteristics** | |
| Time in bed | Mean time (hours) between scored sleep onset and final morning awakening, averaged across all valid main sleep periods. |
| Sleep efficiency | Total sleep time ÷ time in bed × 100, averaged across all valid main sleep periods. |
| Sleep regularity index | Probability that any two epochs 24 hours apart are scored as the same state (sleep vs wake); standardized (z-scored) before regression analysis. |
| **Dementia risk factors** | |
| Hearing impairment | Hearing Handicap Inventory for the Elderly–Screening (HHIE-S) total score ≥ 10.^1^ |
| Hypercholesterolemia | Self-reported history of hypercholesterolemia or use of lipid-lowering medication. |
| Depressive symptoms | Patient-Reported Outcomes Measurement Information System (PROMIS) Depression Short Form 8a v1.0 T-score ≥ 55.^2^ |
| Diabetes | Self-reported history of diabetes or use of antihyperglycemic medication. |
| Current smoking | Self-reported current tobacco use. |
| Hypertension | Self-reported history of hypertension; or measured blood pressure (average of 3 measurements) ≥ 130/80 mmHg; or use of antihypertensive medication. |
| Obesity | Body mass index ≥ 30 kg/m². BMI was calculated from measured height and weight during the in-person interview. |
| Excessive alcohol consumption | Alcohol Use Disorders Identification Test–Concise (AUDIT-C) score ≥ 4 (male) or ≥ 3 (female).^3^ |
| ^1^Ventry IM, Weinstein BE. The Hearing Handicap Inventory for the Elderly: a new tool. Ear Hear. 1982;3(3):128-134. doi:10.1097/00003446-198205000-00006. | |
| ^2^Pilkonis PA, Choi SW, Reise SP, et al. Item banks for measuring emotional distress from the Patient-Reported Outcomes Measurement Information System (PROMIS®): depression, anxiety, and anger. Assessment. 2011;18(3):263-283. doi:10.1177/1073191111411667. | |
| ^3^Bush K, Kivlahan DR, McDonell MB, Fihn SD, Bradley KA. The AUDIT alcohol consumption questions (AUDIT-C): an effective brief screening test for problem drinking. Ambulatory Care Quality Improvement Project (ACQUIP). Alcohol Use Disorders Identification Test. Arch Intern Med. 1998;158(16):1789-1795. doi:10.1001/archinte.158.16.1789. | |

**eTable 2. Characteristics of participants with and without valid accelerometer data**

| **Characteristic** | **Valid accelerometer data N = 854** | **No valid accelerometer data N = 246** | **Absolute SMD** |
| --- | --- | --- | --- |
| **Sociodemographic characteristics** | | | |
| Age, years | 52.8 (8.2); n=854 | 51.6 (8.4); n=246 | 0.14 |
| Female sex | 535/854 (62.6%) | 140/245 (57.1%) | 0.11 |
| Race and ethnicity: Non-Hispanic Black | 340/854 (39.8%) | 115/245 (46.9%) | 0.14 |
| Race and ethnicity: Non-Hispanic White | 330/854 (38.6%) | 80/245 (32.7%) | 0.13 |
| Race and ethnicity: Hispanic or Latino | 114/854 (13.3%) | 36/245 (14.7%) | 0.04 |
| Race and ethnicity: Non-Hispanic Other | 70/854 (8.2%) | 14/245 (5.7%) | 0.10 |
| Education, years | 15.7 (2.7); n=854 | 15.3 (2.8); n=245 | 0.15 |
| Recruitment site: federally qualified health centers | 197/854 (23.1%) | 77/246 (31.3%) | 0.19 |
| **Midlife dementia risk factors** | | | |
| Hearing impairment | 117/847 (13.8%) | 33/228 (14.5%) | 0.02 |
| Hypercholesterolemia | 336/854 (39.3%) | 102/246 (41.5%) | 0.04 |
| Depressive symptoms | 208/844 (24.6%) | 61/227 (26.9%) | 0.05 |
| Diabetes | 182/854 (21.3%) | 55/246 (22.4%) | 0.03 |
| Current smoking | 104/845 (12.3%) | 26/227 (11.5%) | 0.03 |
| Hypertension | 484/854 (56.7%) | 153/246 (62.2%) | 0.11 |
| Obesity | 379/852 (44.5%) | 118/246 (48.0%) | 0.07 |
| Excessive alcohol use | 278/846 (32.9%) | 70/228 (30.7%) | 0.05 |
| **NIH Toolbox Cognition Battery** | | | |
| Fluid Cognition Composite T-score | 48.7 (9.7); n=830 | 45.6 (9.8); n=238 | 0.31 |
| Attention T-score | 47.6 (10.8); n=851 | 45.2 (10.9); n=243 | 0.22 |
| Executive function T-score | 46.8 (8.5); n=850 | 45.6 (8.5); n=243 | 0.14 |
| Episodic memory T-score | 51.0 (10.3); n=838 | 48.6 (9.3); n=240 | 0.25 |
| Working memory T-score | 50.3 (9.5); n=846 | 47.2 (9.5); n=241 | 0.33 |
| Processing speed T-score | 47.9 (9.0); n=851 | 46.5 (9.0); n=243 | 0.16 |
| Crystallized Cognition Composite T-score | 46.0 (13.4); n=845 | 43.8 (13.7); n=242 | 0.16 |
| Receptive language T-score | 48.4 (11.8); n=846 | 46.3 (12.3); n=242 | 0.17 |
| Expressive language T-score | 45.3 (11.7); n=851 | 43.4 (11.8); n=243 | 0.16 |
| Continuous variables are presented as mean (SD); n=available observations. Categorical variables are presented as n/N (%). Cognitive scores were summarized among participants with the corresponding NIH Toolbox outcome available. | | | |
| Absolute standardized mean differences (SMDs) compare participants with and without valid accelerometer data. For continuous variables, the mean difference was divided by the pooled SD; categorical levels were represented as binary indicators. | | | |

**eFigure 1. Correlations among rest–activity rhythm, activity, sleep, and demographic measures**

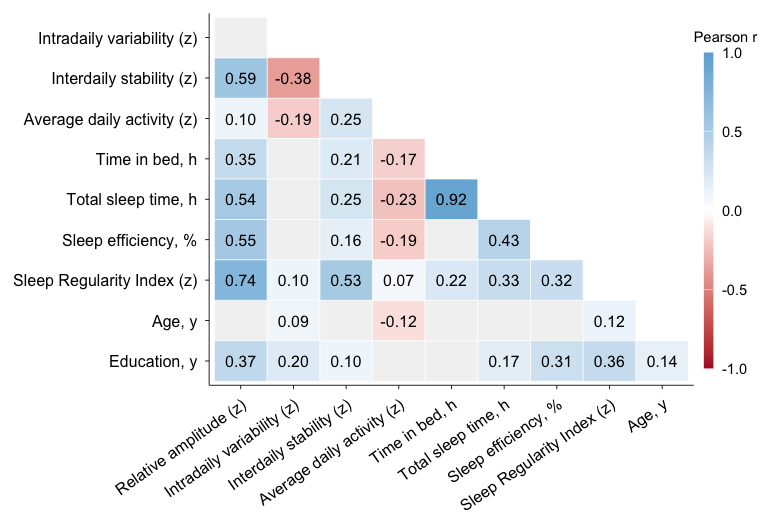

Cells display Pearson correlation coefficients (*r*). Only the lower triangle is shown; cells are blank where *p* ≥ 0.05. Color intensity reflects the magnitude and direction of the correlation (blue, positive; red, negative). Variables labeled '(z)' were standardized to a mean of 0 and an SD of 1 before analysis.

**eTable 3. Associations between relative amplitude and NIH Toolbox Fluid Cognition Composite T-scores**

| **Predictor** | **Model 1 Unadjusted** | **Model 2 RAR + sociodemographic N = 830** | **Model 3 + dementia risk factors N = 817** | **Model 4 + time in bed + sleep efficiency N = 817** | **Model 5 + sleep regularity N = 817** |
| --- | --- | --- | --- | --- | --- |
| Relative amplitude, per SD | **2.68 (2.02, 3.33)** | **1.93 (1.07, 2.79)** | **1.80 (0.92, 2.69)** | **2.27 (1.15, 3.40)** | **1.99 (0.68, 3.29)** |
| Intradaily variability, per SD | **0.95 (0.29, 1.61)** | 0.31 (-0.40, 1.03) | 0.38 (-0.35, 1.11) | 0.35 (-0.38, 1.08) | 0.27 (-0.47, 1.02) |
| Interdaily stability, per SD | **0.86 (0.19, 1.53)** | -0.22 (-1.08, 0.64) | -0.38 (-1.26, 0.50) | -0.51 (-1.41, 0.38) | -0.61 (-1.54, 0.31) |
| Age, years | 0.01 (-0.07, 0.09) | -0.07 (-0.15, 0.01) | -0.02 (-0.11, 0.06) | -0.03 (-0.11, 0.06) | -0.03 (-0.12, 0.06) |
| Female sex | **-1.44 (-2.81, -0.07)** | -1.07 (-2.38, 0.25) | -1.28 (-2.64, 0.08) | -1.10 (-2.48, 0.29) | -1.07 (-2.46, 0.32) |
| Education, years | **1.11 (0.87, 1.34)** | **0.69 (0.41, 0.98)** | **0.64 (0.35, 0.93)** | **0.65 (0.35, 0.94)** | **0.64 (0.34, 0.93)** |
| Recruitment site: FQHC | **-5.72 (-7.26, -4.17)** | **-1.95 (-3.85, -0.05)** | **-2.33 (-4.34, -0.33)** | **-2.35 (-4.37, -0.34)** | **-2.37 (-4.39, -0.35)** |
| Hearing impairment | -1.07 (-2.98, 0.85) |  | -0.62 (-2.43, 1.20) | -0.59 (-2.40, 1.23) | -0.54 (-2.36, 1.27) |
| Hypercholesterolemia | **-1.65 (-3.00, -0.30)** |  | -0.73 (-2.20, 0.73) | -0.66 (-2.13, 0.81) | -0.70 (-2.17, 0.77) |
| Depressive symptoms | -1.44 (-3.01, 0.12) |  | 0.32 (-1.20, 1.84) | 0.25 (-1.27, 1.77) | 0.32 (-1.21, 1.85) |
| Average daily activity, per SD | 0.64 (-0.03, 1.30) |  | **0.76 (0.10, 1.43)** | 0.61 (-0.12, 1.33) | 0.62 (-0.10, 1.35) |
| Diabetes | **-3.57 (-5.13, -2.01)** |  | -0.85 (-2.54, 0.83) | -0.94 (-2.63, 0.75) | -0.95 (-2.64, 0.74) |
| Smoking | **-3.57 (-5.64, -1.51)** |  | 0.21 (-1.97, 2.39) | 0.10 (-2.09, 2.29) | 0.15 (-2.04, 2.34) |
| Hypertension | **-2.00 (-3.34, -0.65)** |  | -0.45 (-1.86, 0.96) | -0.47 (-1.88, 0.94) | -0.44 (-1.85, 0.98) |
| Obesity | **-2.08 (-3.41, -0.75)** |  | 1.05 (-0.35, 2.45) | 1.08 (-0.33, 2.49) | 1.06 (-0.34, 2.47) |
| Excessive alcohol use | **3.06 (1.66, 4.46)** |  | **2.20 (0.85, 3.56)** | **2.20 (0.85, 3.56)** | **2.20 (0.85, 3.56)** |
| Time in bed, h | 0.48 (-0.14, 1.11) |  |  | -0.16 (-0.85, 0.53) | -0.14 (-0.83, 0.55) |
| Sleep efficiency, % | **0.22 (0.10, 0.34)** |  |  | -0.11 (-0.26, 0.04) | -0.10 (-0.25, 0.05) |
| Sleep regularity index, per SD | **2.40 (1.71, 3.08)** |  |  |  | 0.45 (-0.59, 1.50) |
| **Pooled R² (95% CI)** |  | **0.135 (0.094, 0.181)** | **0.156 (0.112, 0.204)** | **0.158 (0.114, 0.206)** | **0.159 (0.115, 0.207)** |
| **Mean BIC** |  | **6073.8** | **6016.2** | **6027.4** | **6033.4** |
| Regression coefficients and variances were combined across 10 completed RAR datasets using Rubin's rules; 95% CIs and two-sided p values were calculated from the pooled estimates. Positive coefficients indicate higher cognitive T-scores. Boldface indicates pooled nominal p < 0.05 and does not denote Benjamini-Hochberg-adjusted significance. For relative amplitude, Benjamini-Hochberg correction was applied across the 8 secondary outcomes separately within each model. RAR, rest-activity rhythm; FQHC, federally qualified health center. | | | | | |
| Model 1 estimates were obtained from separate unadjusted regressions for each predictor, so a common Model 1 R² or BIC is not applicable. Model 2 included intradaily variability, interdaily stability, age, sex, education, and recruitment site type. Model 3 additionally included hearing impairment, hypercholesterolemia, depressive symptoms, average daily activity, diabetes, current smoking, hypertension, obesity, and excessive alcohol use. Model 4 additionally included time in bed and sleep efficiency; Model 5 additionally included the sleep regularity index. | | | | | |
| Reference categories were non-female sex, academic recruitment site, and absence of each binary risk factor. Predictors labeled 'per SD' were standardized (mean = 0, SD = 1). R² and its 95% CI were pooled across the 10 completed datasets using a Fisher z transformation. BIC is the arithmetic mean of the imputation-specific values and is descriptive; it was not combined using Rubin's rules. | | | | | |

**eTable 4. Associations between relative amplitude and NIH Toolbox attention T-scores**

| **Predictor** | **Model 1 Unadjusted** | **Model 2 RAR + sociodemographic N = 851** | **Model 3 + dementia risk factors N = 837** | **Model 4 + time in bed + sleep efficiency N = 837** | **Model 5 + sleep regularity N = 837** |
| --- | --- | --- | --- | --- | --- |
| Relative amplitude, per SD | **2.19 (1.48, 2.91)** | **1.96 (1.00, 2.92)** | **1.95 (0.95, 2.94)** | **2.74 (1.49, 4.00)** | **2.14 (0.66, 3.61)** |
| Intradaily variability, per SD | 0.59 (-0.14, 1.32) | -0.03 (-0.84, 0.78) | -0.01 (-0.84, 0.82) | -0.07 (-0.90, 0.75) | -0.23 (-1.08, 0.62) |
| Interdaily stability, per SD | 0.59 (-0.13, 1.32) | -0.61 (-1.58, 0.36) | -0.70 (-1.70, 0.30) | -0.86 (-1.88, 0.16) | **-1.07 (-2.12, -0.01)** |
| Age, years | **-0.11 (-0.20, -0.02)** | **-0.17 (-0.26, -0.08)** | **-0.15 (-0.25, -0.05)** | **-0.15 (-0.25, -0.05)** | **-0.16 (-0.26, -0.06)** |
| Female sex | **-2.06 (-3.56, -0.56)** | **-1.94 (-3.42, -0.46)** | **-1.99 (-3.54, -0.44)** | **-1.68 (-3.25, -0.10)** | **-1.63 (-3.20, -0.05)** |
| Education, years | **0.81 (0.54, 1.07)** | **0.51 (0.18, 0.83)** | **0.47 (0.14, 0.81)** | **0.46 (0.13, 0.79)** | **0.44 (0.10, 0.77)** |
| Recruitment site: FQHC | **-3.86 (-5.57, -2.15)** | -1.31 (-3.43, 0.81) | -1.49 (-3.77, 0.78) | -1.39 (-3.67, 0.88) | -1.41 (-3.68, 0.87) |
| Hearing impairment | -0.86 (-2.98, 1.25) |  | -0.39 (-2.46, 1.69) | -0.35 (-2.42, 1.73) | -0.28 (-2.36, 1.79) |
| Hypercholesterolemia | -0.83 (-2.32, 0.66) |  | 0.40 (-1.27, 2.06) | 0.47 (-1.19, 2.14) | 0.40 (-1.27, 2.07) |
| Depressive symptoms | -0.82 (-2.51, 0.88) |  | 0.49 (-1.22, 2.21) | 0.44 (-1.27, 2.16) | 0.58 (-1.14, 2.31) |
| Average daily activity, per SD | 0.27 (-0.46, 0.99) |  | 0.28 (-0.48, 1.04) | -0.08 (-0.90, 0.75) | -0.04 (-0.86, 0.79) |
| Diabetes | **-2.65 (-4.37, -0.92)** |  | -0.74 (-2.67, 1.18) | -0.94 (-2.87, 0.99) | -0.98 (-2.91, 0.95) |
| Smoking | **-2.96 (-5.18, -0.74)** |  | -0.09 (-2.54, 2.37) | -0.33 (-2.79, 2.13) | -0.20 (-2.67, 2.26) |
| Hypertension | **-2.03 (-3.50, -0.55)** |  | -0.71 (-2.32, 0.90) | -0.72 (-2.32, 0.89) | -0.64 (-2.25, 0.97) |
| Obesity | -1.20 (-2.67, 0.26) |  | 0.96 (-0.64, 2.56) | 0.87 (-0.73, 2.48) | 0.83 (-0.77, 2.44) |
| Excessive alcohol use | **1.94 (0.39, 3.50)** |  | 1.13 (-0.41, 2.68) | 1.11 (-0.44, 2.65) | 1.10 (-0.44, 2.64) |
| Time in bed, h | 0.01 (-0.66, 0.69) |  |  | -0.72 (-1.48, 0.04) | -0.68 (-1.44, 0.08) |
| Sleep efficiency, % | **0.20 (0.07, 0.33)** |  |  | -0.14 (-0.31, 0.03) | -0.12 (-0.29, 0.06) |
| Sleep regularity index, per SD | **2.04 (1.32, 2.75)** |  |  |  | 0.90 (-0.26, 2.06) |
| **Pooled R² (95% CI)** |  | **0.086 (0.053, 0.125)** | **0.090 (0.056, 0.130)** | **0.095 (0.061, 0.136)** | **0.098 (0.063, 0.139)** |
| **Mean BIC** |  | **6450.6** | **6399.6** | **6408.2** | **6412.6** |
| Regression coefficients and variances were combined across 10 completed RAR datasets using Rubin's rules; 95% CIs and two-sided p values were calculated from the pooled estimates. Positive coefficients indicate higher cognitive T-scores. Boldface indicates pooled nominal p < 0.05 and does not denote Benjamini-Hochberg-adjusted significance. For relative amplitude, Benjamini-Hochberg correction was applied across the 8 secondary outcomes separately within each model. RAR, rest-activity rhythm; FQHC, federally qualified health center. | | | | | |
| Model 1 estimates were obtained from separate unadjusted regressions for each predictor, so a common Model 1 R² or BIC is not applicable. Model 2 included intradaily variability, interdaily stability, age, sex, education, and recruitment site type. Model 3 additionally included hearing impairment, hypercholesterolemia, depressive symptoms, average daily activity, diabetes, current smoking, hypertension, obesity, and excessive alcohol use. Model 4 additionally included time in bed and sleep efficiency; Model 5 additionally included the sleep regularity index. | | | | | |
| Reference categories were non-female sex, academic recruitment site, and absence of each binary risk factor. Predictors labeled 'per SD' were standardized (mean = 0, SD = 1). R² and its 95% CI were pooled across the 10 completed datasets using a Fisher z transformation. BIC is the arithmetic mean of the imputation-specific values and is descriptive; it was not combined using Rubin's rules. | | | | | |

**eTable 5. Associations between relative amplitude and NIH Toolbox executive function T-scores**

| **Predictor** | **Model 1 Unadjusted** | **Model 2 RAR + sociodemographic N = 850** | **Model 3 + dementia risk factors N = 836** | **Model 4 + time in bed + sleep efficiency N = 836** | **Model 5 + sleep regularity N = 836** |
| --- | --- | --- | --- | --- | --- |
| Relative amplitude, per SD | **1.88 (1.32, 2.45)** | **1.33 (0.57, 2.08)** | **1.39 (0.62, 2.16)** | **1.49 (0.51, 2.47)** | **1.23 (0.09, 2.38)** |
| Intradaily variability, per SD | **0.81 (0.24, 1.38)** | 0.44 (-0.19, 1.06) | 0.49 (-0.15, 1.13) | 0.48 (-0.16, 1.12) | 0.41 (-0.25, 1.07) |
| Interdaily stability, per SD | **0.67 (0.10, 1.24)** | 0.01 (-0.74, 0.76) | -0.08 (-0.86, 0.69) | -0.10 (-0.90, 0.69) | -0.19 (-1.01, 0.62) |
| Age, years | -0.04 (-0.11, 0.03) | **-0.09 (-0.16, -0.02)** | -0.05 (-0.13, 0.03) | -0.05 (-0.13, 0.02) | -0.05 (-0.13, 0.02) |
| Female sex | **-1.81 (-2.99, -0.64)** | **-1.62 (-2.77, -0.47)** | **-1.89 (-3.08, -0.69)** | **-1.85 (-3.07, -0.63)** | **-1.83 (-3.05, -0.60)** |
| Education, years | **0.78 (0.57, 0.98)** | **0.55 (0.30, 0.80)** | **0.54 (0.28, 0.80)** | **0.54 (0.28, 0.80)** | **0.53 (0.27, 0.79)** |
| Recruitment site: FQHC | **-3.36 (-4.70, -2.02)** | -0.43 (-2.08, 1.21) | -1.17 (-2.93, 0.58) | -1.16 (-2.93, 0.60) | -1.17 (-2.93, 0.60) |
| Hearing impairment | -0.64 (-2.30, 1.01) |  | -0.32 (-1.92, 1.28) | -0.31 (-1.92, 1.29) | -0.28 (-1.89, 1.32) |
| Hypercholesterolemia | **-1.20 (-2.37, -0.04)** |  | -0.70 (-1.99, 0.59) | -0.69 (-1.98, 0.60) | -0.73 (-2.02, 0.57) |
| Depressive symptoms | -0.50 (-1.83, 0.84) |  | 0.79 (-0.54, 2.11) | 0.78 (-0.55, 2.11) | 0.84 (-0.50, 2.17) |
| Average daily activity, per SD | 0.47 (-0.10, 1.04) |  | 0.50 (-0.08, 1.09) | 0.46 (-0.18, 1.10) | 0.48 (-0.16, 1.12) |
| Diabetes | **-2.16 (-3.51, -0.81)** |  | 0.05 (-1.44, 1.54) | 0.03 (-1.47, 1.53) | 0.01 (-1.49, 1.51) |
| Smoking | -1.65 (-3.40, 0.09) |  | 1.22 (-0.68, 3.12) | 1.19 (-0.72, 3.10) | 1.24 (-0.67, 3.15) |
| Hypertension | **-1.71 (-2.86, -0.55)** |  | -0.66 (-1.90, 0.59) | -0.66 (-1.90, 0.59) | -0.63 (-1.87, 0.62) |
| Obesity | -0.90 (-2.05, 0.24) |  | **1.37 (0.13, 2.60)** | **1.36 (0.12, 2.61)** | **1.34 (0.10, 2.59)** |
| Excessive alcohol use | **1.99 (0.78, 3.20)** |  | **1.39 (0.19, 2.58)** | **1.38 (0.19, 2.58)** | **1.38 (0.18, 2.58)** |
| Time in bed, h | 0.21 (-0.32, 0.73) |  |  | -0.08 (-0.67, 0.51) | -0.06 (-0.66, 0.53) |
| Sleep efficiency, % | **0.18 (0.08, 0.28)** |  |  | -0.02 (-0.15, 0.11) | -0.01 (-0.14, 0.13) |
| Sleep regularity index, per SD | **1.70 (1.13, 2.27)** |  |  |  | 0.39 (-0.51, 1.30) |
| **Pooled R² (95% CI)** |  | **0.100 (0.065, 0.141)** | **0.115 (0.078, 0.159)** | **0.116 (0.078, 0.159)** | **0.116 (0.078, 0.160)** |
| **Mean BIC** |  | **6013.0** | **5960.0** | **5973.4** | **5979.4** |
| Regression coefficients and variances were combined across 10 completed RAR datasets using Rubin's rules; 95% CIs and two-sided p values were calculated from the pooled estimates. Positive coefficients indicate higher cognitive T-scores. Boldface indicates pooled nominal p < 0.05 and does not denote Benjamini-Hochberg-adjusted significance. For relative amplitude, Benjamini-Hochberg correction was applied across the 8 secondary outcomes separately within each model. RAR, rest-activity rhythm; FQHC, federally qualified health center. | | | | | |
| Model 1 estimates were obtained from separate unadjusted regressions for each predictor, so a common Model 1 R² or BIC is not applicable. Model 2 included intradaily variability, interdaily stability, age, sex, education, and recruitment site type. Model 3 additionally included hearing impairment, hypercholesterolemia, depressive symptoms, average daily activity, diabetes, current smoking, hypertension, obesity, and excessive alcohol use. Model 4 additionally included time in bed and sleep efficiency; Model 5 additionally included the sleep regularity index. | | | | | |
| Reference categories were non-female sex, academic recruitment site, and absence of each binary risk factor. Predictors labeled 'per SD' were standardized (mean = 0, SD = 1). R² and its 95% CI were pooled across the 10 completed datasets using a Fisher z transformation. BIC is the arithmetic mean of the imputation-specific values and is descriptive; it was not combined using Rubin's rules. | | | | | |

**eTable 6. Associations between relative amplitude and NIH Toolbox episodic memory T-scores**

| **Predictor** | **Model 1 Unadjusted** | **Model 2 RAR + sociodemographic N = 838** | **Model 3 + dementia risk factors N = 825** | **Model 4 + time in bed + sleep efficiency N = 825** | **Model 5 + sleep regularity N = 825** |
| --- | --- | --- | --- | --- | --- |
| Relative amplitude, per SD | **2.29 (1.60, 2.98)** | **1.92 (0.99, 2.85)** | **1.76 (0.80, 2.72)** | **2.03 (0.81, 3.25)** | **2.28 (0.85, 3.71)** |
| Intradaily variability, per SD | 0.29 (-0.41, 0.98) | -0.26 (-1.04, 0.51) | -0.13 (-0.92, 0.66) | -0.15 (-0.94, 0.65) | -0.08 (-0.90, 0.74) |
| Interdaily stability, per SD | **0.78 (0.08, 1.48)** | -0.59 (-1.52, 0.35) | -0.67 (-1.63, 0.29) | -0.75 (-1.73, 0.23) | -0.67 (-1.68, 0.35) |
| Age, years | 0.01 (-0.07, 0.10) | -0.05 (-0.14, 0.03) | -0.05 (-0.15, 0.04) | -0.05 (-0.15, 0.04) | -0.05 (-0.14, 0.05) |
| Female sex | 0.16 (-1.28, 1.60) | 0.36 (-1.06, 1.78) | 0.40 (-1.09, 1.88) | 0.50 (-1.01, 2.02) | 0.49 (-1.03, 2.00) |
| Education, years | **0.79 (0.53, 1.04)** | **0.37 (0.06, 0.69)** | **0.37 (0.05, 0.69)** | **0.38 (0.05, 0.70)** | **0.38 (0.06, 0.71)** |
| Recruitment site: FQHC | **-4.78 (-6.40, -3.16)** | **-2.59 (-4.64, -0.54)** | **-2.66 (-4.85, -0.47)** | **-2.68 (-4.88, -0.48)** | **-2.68 (-4.88, -0.47)** |
| Hearing impairment | 1.19 (-0.82, 3.20) |  | 1.44 (-0.55, 3.42) | 1.45 (-0.53, 3.44) | 1.42 (-0.56, 3.41) |
| Hypercholesterolemia | -0.57 (-1.99, 0.85) |  | 0.62 (-0.97, 2.22) | 0.67 (-0.93, 2.27) | 0.70 (-0.91, 2.30) |
| Depressive symptoms | -1.60 (-3.23, 0.04) |  | -0.33 (-1.99, 1.33) | -0.36 (-2.03, 1.30) | -0.42 (-2.10, 1.25) |
| Average daily activity, per SD | 0.51 (-0.19, 1.21) |  | 0.65 (-0.08, 1.38) | 0.56 (-0.23, 1.35) | 0.55 (-0.25, 1.34) |
| Diabetes | **-2.63 (-4.27, -0.99)** |  | -0.98 (-2.82, 0.86) | -1.03 (-2.88, 0.82) | -1.01 (-2.86, 0.84) |
| Smoking | **-2.84 (-4.98, -0.70)** |  | 0.10 (-2.27, 2.47) | 0.04 (-2.34, 2.42) | -0.01 (-2.39, 2.37) |
| Hypertension | **-1.61 (-3.02, -0.19)** |  | -0.33 (-1.88, 1.21) | -0.34 (-1.89, 1.20) | -0.37 (-1.92, 1.17) |
| Obesity | **-1.66 (-3.06, -0.27)** |  | 0.74 (-0.79, 2.27) | 0.76 (-0.78, 2.30) | 0.78 (-0.76, 2.32) |
| Excessive alcohol use | **2.30 (0.82, 3.78)** |  | **1.79 (0.31, 3.27)** | **1.79 (0.31, 3.28)** | **1.80 (0.32, 3.28)** |
| Time in bed, h | 0.55 (-0.11, 1.20) |  |  | -0.06 (-0.82, 0.69) | -0.09 (-0.84, 0.67) |
| Sleep efficiency, % | **0.23 (0.10, 0.35)** |  |  | -0.07 (-0.23, 0.10) | -0.08 (-0.24, 0.09) |
| Sleep regularity index, per SD | **1.59 (0.88, 2.29)** |  |  |  | -0.37 (-1.50, 0.75) |
| **Pooled R² (95% CI)** |  | **0.077 (0.045, 0.114)** | **0.089 (0.055, 0.129)** | **0.090 (0.056, 0.130)** | **0.090 (0.056, 0.131)** |
| **Mean BIC** |  | **6271.7** | **6227.9** | **6240.7** | **6247.0** |
| Regression coefficients and variances were combined across 10 completed RAR datasets using Rubin's rules; 95% CIs and two-sided p values were calculated from the pooled estimates. Positive coefficients indicate higher cognitive T-scores. Boldface indicates pooled nominal p < 0.05 and does not denote Benjamini-Hochberg-adjusted significance. For relative amplitude, Benjamini-Hochberg correction was applied across the 8 secondary outcomes separately within each model. RAR, rest-activity rhythm; FQHC, federally qualified health center. | | | | | |
| Model 1 estimates were obtained from separate unadjusted regressions for each predictor, so a common Model 1 R² or BIC is not applicable. Model 2 included intradaily variability, interdaily stability, age, sex, education, and recruitment site type. Model 3 additionally included hearing impairment, hypercholesterolemia, depressive symptoms, average daily activity, diabetes, current smoking, hypertension, obesity, and excessive alcohol use. Model 4 additionally included time in bed and sleep efficiency; Model 5 additionally included the sleep regularity index. | | | | | |
| Reference categories were non-female sex, academic recruitment site, and absence of each binary risk factor. Predictors labeled 'per SD' were standardized (mean = 0, SD = 1). R² and its 95% CI were pooled across the 10 completed datasets using a Fisher z transformation. BIC is the arithmetic mean of the imputation-specific values and is descriptive; it was not combined using Rubin's rules. | | | | | |

**eTable 7. Associations between relative amplitude and NIH Toolbox working memory T-scores**

| **Predictor** | **Model 1 Unadjusted** | **Model 2 RAR + sociodemographic N = 846** | **Model 3 + dementia risk factors N = 832** | **Model 4 + time in bed + sleep efficiency N = 832** | **Model 5 + sleep regularity N = 832** |
| --- | --- | --- | --- | --- | --- |
| Relative amplitude, per SD | **1.83 (1.21, 2.46)** | 0.62 (-0.20, 1.44) | 0.28 (-0.56, 1.11) | 0.45 (-0.61, 1.50) | 0.40 (-0.84, 1.64) |
| Intradaily variability, per SD | **1.41 (0.78, 2.04)** | 0.66 (-0.03, 1.34) | 0.62 (-0.07, 1.31) | 0.61 (-0.08, 1.30) | 0.60 (-0.12, 1.31) |
| Interdaily stability, per SD | 0.47 (-0.17, 1.10) | 0.22 (-0.60, 1.04) | 0.08 (-0.76, 0.92) | 0.01 (-0.84, 0.87) | -0.00 (-0.89, 0.88) |
| Age, years | **0.08 (0.00, 0.16)** | -0.02 (-0.09, 0.06) | 0.03 (-0.06, 0.11) | 0.03 (-0.06, 0.11) | 0.02 (-0.06, 0.11) |
| Female sex | **-2.24 (-3.55, -0.93)** | **-1.56 (-2.82, -0.31)** | **-1.83 (-3.13, -0.53)** | **-1.76 (-3.09, -0.44)** | **-1.76 (-3.09, -0.43)** |
| Education, years | **1.16 (0.94, 1.38)** | **0.75 (0.47, 1.02)** | **0.67 (0.39, 0.95)** | **0.68 (0.39, 0.96)** | **0.67 (0.39, 0.96)** |
| Recruitment site: FQHC | **-6.73 (-8.18, -5.28)** | **-3.06 (-4.86, -1.26)** | **-2.82 (-4.73, -0.91)** | **-2.87 (-4.79, -0.95)** | **-2.87 (-4.79, -0.95)** |
| Hearing impairment | -1.71 (-3.55, 0.14) |  | -1.32 (-3.06, 0.42) | -1.31 (-3.05, 0.43) | -1.30 (-3.04, 0.44) |
| Hypercholesterolemia | **-1.45 (-2.75, -0.15)** |  | -1.12 (-2.51, 0.28) | -1.08 (-2.48, 0.31) | -1.09 (-2.49, 0.31) |
| Depressive symptoms | **-3.65 (-5.13, -2.17)** |  | **-1.81 (-3.25, -0.37)** | **-1.84 (-3.29, -0.40)** | **-1.83 (-3.28, -0.38)** |
| Average daily activity, per SD | 0.28 (-0.36, 0.92) |  | 0.53 (-0.11, 1.16) | 0.48 (-0.21, 1.18) | 0.49 (-0.21, 1.18) |
| Diabetes | **-2.83 (-4.34, -1.32)** |  | -0.64 (-2.25, 0.97) | -0.66 (-2.28, 0.96) | -0.67 (-2.29, 0.95) |
| Smoking | **-5.22 (-7.16, -3.29)** |  | -1.16 (-3.22, 0.91) | -1.19 (-3.27, 0.88) | -1.18 (-3.26, 0.90) |
| Hypertension | **-1.89 (-3.18, -0.59)** |  | -0.33 (-1.68, 1.02) | -0.34 (-1.69, 1.02) | -0.33 (-1.68, 1.02) |
| Obesity | **-2.54 (-3.82, -1.27)** |  | 0.28 (-1.06, 1.62) | 0.33 (-1.02, 1.68) | 0.33 (-1.03, 1.68) |
| Excessive alcohol use | **2.47 (1.12, 3.83)** |  | **2.05 (0.76, 3.35)** | **2.06 (0.76, 3.36)** | **2.06 (0.76, 3.36)** |
| Time in bed, h | 0.26 (-0.33, 0.85) |  |  | 0.05 (-0.59, 0.69) | 0.05 (-0.59, 0.69) |
| Sleep efficiency, % | **0.18 (0.07, 0.30)** |  |  | -0.05 (-0.20, 0.09) | -0.05 (-0.20, 0.09) |
| Sleep regularity index, per SD | **1.85 (1.22, 2.49)** |  |  |  | 0.07 (-0.91, 1.06) |
| **Pooled R² (95% CI)** |  | **0.145 (0.103, 0.191)** | **0.174 (0.129, 0.222)** | **0.175 (0.130, 0.223)** | **0.175 (0.130, 0.223)** |
| **Mean BIC** |  | **6129.4** | **6064.2** | **6077.0** | **6083.7** |
| Regression coefficients and variances were combined across 10 completed RAR datasets using Rubin's rules; 95% CIs and two-sided p values were calculated from the pooled estimates. Positive coefficients indicate higher cognitive T-scores. Boldface indicates pooled nominal p < 0.05 and does not denote Benjamini-Hochberg-adjusted significance. For relative amplitude, Benjamini-Hochberg correction was applied across the 8 secondary outcomes separately within each model. RAR, rest-activity rhythm; FQHC, federally qualified health center. | | | | | |
| Model 1 estimates were obtained from separate unadjusted regressions for each predictor, so a common Model 1 R² or BIC is not applicable. Model 2 included intradaily variability, interdaily stability, age, sex, education, and recruitment site type. Model 3 additionally included hearing impairment, hypercholesterolemia, depressive symptoms, average daily activity, diabetes, current smoking, hypertension, obesity, and excessive alcohol use. Model 4 additionally included time in bed and sleep efficiency; Model 5 additionally included the sleep regularity index. | | | | | |
| Reference categories were non-female sex, academic recruitment site, and absence of each binary risk factor. Predictors labeled 'per SD' were standardized (mean = 0, SD = 1). R² and its 95% CI were pooled across the 10 completed datasets using a Fisher z transformation. BIC is the arithmetic mean of the imputation-specific values and is descriptive; it was not combined using Rubin's rules. | | | | | |

**eTable 8. Associations between relative amplitude and NIH Toolbox processing speed T-scores**

| **Predictor** | **Model 1 Unadjusted** | **Model 2 RAR + sociodemographic N = 851** | **Model 3 + dementia risk factors N = 837** | **Model 4 + time in bed + sleep efficiency N = 837** | **Model 5 + sleep regularity N = 837** |
| --- | --- | --- | --- | --- | --- |
| Relative amplitude, per SD | **1.29 (0.69, 1.89)** | **1.15 (0.32, 1.98)** | **1.12 (0.27, 1.97)** | **1.56 (0.49, 2.64)** | 0.92 (-0.34, 2.18) |
| Intradaily variability, per SD | 0.20 (-0.41, 0.81) | 0.14 (-0.55, 0.84) | 0.14 (-0.56, 0.85) | 0.11 (-0.60, 0.82) | -0.06 (-0.78, 0.67) |
| Interdaily stability, per SD | 0.57 (-0.04, 1.17) | -0.17 (-1.00, 0.67) | -0.34 (-1.20, 0.51) | -0.48 (-1.35, 0.39) | -0.70 (-1.60, 0.20) |
| Age, years | -0.05 (-0.13, 0.02) | -0.07 (-0.14, 0.01) | -0.03 (-0.12, 0.05) | -0.03 (-0.12, 0.05) | -0.04 (-0.12, 0.04) |
| Female sex | 0.37 (-0.89, 1.63) | 0.35 (-0.92, 1.63) | 0.30 (-1.02, 1.62) | 0.47 (-0.87, 1.82) | 0.52 (-0.82, 1.87) |
| Education, years | **0.41 (0.19, 0.64)** | **0.33 (0.05, 0.61)** | **0.29 (0.01, 0.58)** | **0.30 (0.02, 0.59)** | 0.28 (-0.01, 0.57) |
| Recruitment site: FQHC | -1.15 (-2.59, 0.30) | 0.56 (-1.27, 2.38) | 0.08 (-1.86, 2.02) | 0.04 (-1.91, 1.99) | 0.03 (-1.92, 1.97) |
| Hearing impairment | -0.92 (-2.69, 0.85) |  | -0.57 (-2.35, 1.20) | -0.55 (-2.32, 1.23) | -0.48 (-2.25, 1.29) |
| Hypercholesterolemia | **-1.61 (-2.85, -0.37)** |  | -0.72 (-2.14, 0.70) | -0.66 (-2.08, 0.77) | -0.74 (-2.16, 0.69) |
| Depressive symptoms | 0.08 (-1.34, 1.49) |  | 0.76 (-0.71, 2.22) | 0.70 (-0.77, 2.17) | 0.85 (-0.62, 2.32) |
| Average daily activity, per SD | 0.57 (-0.04, 1.18) |  | 0.47 (-0.18, 1.12) | 0.32 (-0.39, 1.02) | 0.36 (-0.35, 1.07) |
| Diabetes | **-2.03 (-3.48, -0.59)** |  | -0.41 (-2.05, 1.24) | -0.49 (-2.14, 1.16) | -0.54 (-2.19, 1.11) |
| Smoking | -0.73 (-2.59, 1.13) |  | 0.56 (-1.54, 2.65) | 0.45 (-1.66, 2.55) | 0.58 (-1.52, 2.69) |
| Hypertension | **-1.27 (-2.51, -0.03)** |  | -0.37 (-1.75, 1.00) | -0.38 (-1.76, 0.99) | -0.30 (-1.68, 1.07) |
| Obesity | -0.81 (-2.04, 0.42) |  | 0.34 (-1.03, 1.70) | 0.38 (-0.99, 1.76) | 0.34 (-1.03, 1.71) |
| Excessive alcohol use | **1.90 (0.61, 3.19)** |  | 1.18 (-0.14, 2.50) | 1.19 (-0.13, 2.51) | 1.18 (-0.14, 2.50) |
| Time in bed, h | 0.29 (-0.27, 0.85) |  |  | -0.10 (-0.75, 0.55) | -0.05 (-0.70, 0.60) |
| Sleep efficiency, % | 0.07 (-0.04, 0.18) |  |  | -0.11 (-0.26, 0.03) | -0.09 (-0.24, 0.06) |
| Sleep regularity index, per SD | **1.31 (0.71, 1.91)** |  |  |  | 0.95 (-0.04, 1.95) |
| **Pooled R² (95% CI)** |  | **0.031 (0.012, 0.058)** | **0.043 (0.020, 0.074)** | **0.046 (0.022, 0.077)** | **0.050 (0.025, 0.082)** |
| **Mean BIC** |  | **6195.7** | **6135.4** | **6146.5** | **6149.6** |
| Regression coefficients and variances were combined across 10 completed RAR datasets using Rubin's rules; 95% CIs and two-sided p values were calculated from the pooled estimates. Positive coefficients indicate higher cognitive T-scores. Boldface indicates pooled nominal p < 0.05 and does not denote Benjamini-Hochberg-adjusted significance. For relative amplitude, Benjamini-Hochberg correction was applied across the 8 secondary outcomes separately within each model. RAR, rest-activity rhythm; FQHC, federally qualified health center. | | | | | |
| Model 1 estimates were obtained from separate unadjusted regressions for each predictor, so a common Model 1 R² or BIC is not applicable. Model 2 included intradaily variability, interdaily stability, age, sex, education, and recruitment site type. Model 3 additionally included hearing impairment, hypercholesterolemia, depressive symptoms, average daily activity, diabetes, current smoking, hypertension, obesity, and excessive alcohol use. Model 4 additionally included time in bed and sleep efficiency; Model 5 additionally included the sleep regularity index. | | | | | |
| Reference categories were non-female sex, academic recruitment site, and absence of each binary risk factor. Predictors labeled 'per SD' were standardized (mean = 0, SD = 1). R² and its 95% CI were pooled across the 10 completed datasets using a Fisher z transformation. BIC is the arithmetic mean of the imputation-specific values and is descriptive; it was not combined using Rubin's rules. | | | | | |

**eTable 9. Associations between relative amplitude and NIH Toolbox Crystallized Cognition Composite T-scores**

| **Predictor** | **Model 1 Unadjusted** | **Model 2 RAR + sociodemographic N = 845** | **Model 3 + dementia risk factors N = 832** | **Model 4 + time in bed + sleep efficiency N = 832** | **Model 5 + sleep regularity N = 832** |
| --- | --- | --- | --- | --- | --- |
| Relative amplitude, per SD | **3.80 (2.91, 4.68)** | **1.23 (0.17, 2.28)** | 1.00 (-0.08, 2.09) | 0.71 (-0.66, 2.08) | 0.89 (-0.71, 2.50) |
| Intradaily variability, per SD | **2.32 (1.43, 3.22)** | 0.68 (-0.20, 1.56) | 0.84 (-0.06, 1.73) | 0.86 (-0.04, 1.76) | 0.90 (-0.02, 1.83) |
| Interdaily stability, per SD | **0.97 (0.06, 1.88)** | 0.13 (-0.93, 1.18) | 0.21 (-0.88, 1.29) | 0.28 (-0.83, 1.39) | 0.34 (-0.80, 1.49) |
| Age, years | 0.10 (-0.01, 0.22) | **-0.10 (-0.20, -0.00)** | -0.06 (-0.16, 0.05) | -0.05 (-0.16, 0.05) | -0.05 (-0.16, 0.06) |
| Female sex | **-3.09 (-4.95, -1.23)** | **-1.93 (-3.55, -0.32)** | **-2.10 (-3.77, -0.42)** | **-2.21 (-3.91, -0.50)** | **-2.22 (-3.93, -0.51)** |
| Education, years | **2.41 (2.12, 2.70)** | **1.57 (1.22, 1.92)** | **1.53 (1.17, 1.89)** | **1.53 (1.17, 1.89)** | **1.54 (1.17, 1.90)** |
| Recruitment site: FQHC | **-14.16 (-16.09, -12.24)** | **-7.42 (-9.73, -5.10)** | **-7.59 (-10.05, -5.13)** | **-7.59 (-10.06, -5.12)** | **-7.59 (-10.06, -5.12)** |
| Hearing impairment | -0.89 (-3.52, 1.73) |  | -0.94 (-3.18, 1.30) | -0.95 (-3.20, 1.29) | -0.97 (-3.22, 1.27) |
| Hypercholesterolemia | **-1.96 (-3.81, -0.11)** |  | -0.57 (-2.37, 1.24) | -0.60 (-2.41, 1.20) | -0.58 (-2.40, 1.23) |
| Depressive symptoms | **-2.13 (-4.24, -0.02)** |  | 1.58 (-0.27, 3.44) | 1.61 (-0.25, 3.47) | 1.57 (-0.30, 3.44) |
| Average daily activity, per SD | -0.06 (-0.96, 0.85) |  | 0.47 (-0.35, 1.29) | 0.58 (-0.31, 1.47) | 0.57 (-0.33, 1.46) |
| Diabetes | **-6.30 (-8.42, -4.19)** |  | **-2.42 (-4.50, -0.33)** | **-2.36 (-4.45, -0.26)** | **-2.34 (-4.44, -0.25)** |
| Smoking | **-8.25 (-10.96, -5.54)** |  | 0.31 (-2.35, 2.96) | 0.38 (-2.28, 3.05) | 0.34 (-2.33, 3.02) |
| Hypertension | **-2.95 (-4.79, -1.11)** |  | 0.30 (-1.44, 2.05) | 0.31 (-1.43, 2.06) | 0.29 (-1.46, 2.04) |
| Obesity | **-4.85 (-6.65, -3.05)** |  | 0.41 (-1.32, 2.14) | 0.41 (-1.33, 2.15) | 0.42 (-1.32, 2.16) |
| Excessive alcohol use | **3.51 (1.59, 5.43)** |  | **2.30 (0.63, 3.97)** | **2.30 (0.63, 3.98)** | **2.31 (0.63, 3.98)** |
| Time in bed, h | 0.61 (-0.23, 1.45) |  |  | 0.15 (-0.67, 0.98) | 0.14 (-0.69, 0.97) |
| Sleep efficiency, % | **0.50 (0.34, 0.66)** |  |  | 0.06 (-0.12, 0.25) | 0.06 (-0.13, 0.24) |
| Sleep regularity index, per SD | **3.25 (2.36, 4.15)** |  |  |  | -0.27 (-1.54, 0.99) |
| **Pooled R² (95% CI)** |  | **0.299 (0.247, 0.351)** | **0.313 (0.261, 0.365)** | **0.313 (0.261, 0.366)** | **0.313 (0.262, 0.366)** |
| **Mean BIC** |  | **6547.3** | **6488.4** | **6501.3** | **6507.9** |
| Regression coefficients and variances were combined across 10 completed RAR datasets using Rubin's rules; 95% CIs and two-sided p values were calculated from the pooled estimates. Positive coefficients indicate higher cognitive T-scores. Boldface indicates pooled nominal p < 0.05 and does not denote Benjamini-Hochberg-adjusted significance. For relative amplitude, Benjamini-Hochberg correction was applied across the 8 secondary outcomes separately within each model. RAR, rest-activity rhythm; FQHC, federally qualified health center. | | | | | |
| Model 1 estimates were obtained from separate unadjusted regressions for each predictor, so a common Model 1 R² or BIC is not applicable. Model 2 included intradaily variability, interdaily stability, age, sex, education, and recruitment site type. Model 3 additionally included hearing impairment, hypercholesterolemia, depressive symptoms, average daily activity, diabetes, current smoking, hypertension, obesity, and excessive alcohol use. Model 4 additionally included time in bed and sleep efficiency; Model 5 additionally included the sleep regularity index. | | | | | |
| Reference categories were non-female sex, academic recruitment site, and absence of each binary risk factor. Predictors labeled 'per SD' were standardized (mean = 0, SD = 1). R² and its 95% CI were pooled across the 10 completed datasets using a Fisher z transformation. BIC is the arithmetic mean of the imputation-specific values and is descriptive; it was not combined using Rubin's rules. | | | | | |

**eTable 10. Associations between relative amplitude and NIH Toolbox receptive language T-scores**

| **Predictor** | **Model 1 Unadjusted** | **Model 2 RAR + sociodemographic N = 846** | **Model 3 + dementia risk factors N = 833** | **Model 4 + time in bed + sleep efficiency N = 833** | **Model 5 + sleep regularity N = 833** |
| --- | --- | --- | --- | --- | --- |
| Relative amplitude, per SD | **3.21 (2.43, 3.98)** | **1.13 (0.20, 2.07)** | 0.86 (-0.11, 1.82) | 0.59 (-0.63, 1.80) | 0.70 (-0.73, 2.12) |
| Intradaily variability, per SD | **1.92 (1.13, 2.70)** | 0.37 (-0.42, 1.15) | 0.47 (-0.33, 1.26) | 0.49 (-0.31, 1.29) | 0.52 (-0.31, 1.34) |
| Interdaily stability, per SD | 0.71 (-0.09, 1.51) | -0.08 (-1.01, 0.86) | -0.05 (-1.01, 0.91) | 0.02 (-0.97, 1.00) | 0.06 (-0.96, 1.07) |
| Age, years | **0.13 (0.03, 0.23)** | -0.04 (-0.13, 0.04) | -0.00 (-0.10, 0.09) | -0.00 (-0.09, 0.09) | 0.00 (-0.09, 0.10) |
| Female sex | **-3.29 (-4.91, -1.66)** | **-2.28 (-3.71, -0.85)** | **-2.41 (-3.89, -0.92)** | **-2.51 (-4.03, -1.00)** | **-2.52 (-4.04, -1.00)** |
| Education, years | **2.04 (1.78, 2.29)** | **1.29 (0.98, 1.61)** | **1.25 (0.93, 1.57)** | **1.25 (0.93, 1.57)** | **1.26 (0.93, 1.58)** |
| Recruitment site: FQHC | **-12.34 (-14.03, -10.64)** | **-6.51 (-8.57, -4.46)** | **-6.58 (-8.76, -4.39)** | **-6.58 (-8.78, -4.38)** | **-6.58 (-8.78, -4.38)** |
| Hearing impairment | -0.50 (-2.80, 1.81) |  | -0.68 (-2.67, 1.31) | -0.70 (-2.69, 1.30) | -0.71 (-2.71, 1.29) |
| Hypercholesterolemia | -1.28 (-2.91, 0.34) |  | -0.29 (-1.89, 1.31) | -0.32 (-1.93, 1.28) | -0.31 (-1.92, 1.29) |
| Depressive symptoms | **-2.23 (-4.08, -0.37)** |  | 0.95 (-0.70, 2.60) | 0.98 (-0.68, 2.63) | 0.95 (-0.71, 2.62) |
| Average daily activity, per SD | -0.07 (-0.87, 0.72) |  | 0.40 (-0.33, 1.12) | 0.50 (-0.29, 1.29) | 0.49 (-0.30, 1.29) |
| Diabetes | **-5.38 (-7.24, -3.52)** |  | **-2.11 (-3.96, -0.26)** | **-2.06 (-3.91, -0.20)** | **-2.05 (-3.91, -0.19)** |
| Smoking | **-7.25 (-9.63, -4.86)** |  | 0.14 (-2.21, 2.50) | 0.22 (-2.15, 2.58) | 0.19 (-2.19, 2.57) |
| Hypertension | **-2.84 (-4.45, -1.23)** |  | -0.42 (-1.97, 1.12) | -0.42 (-1.96, 1.13) | -0.43 (-1.98, 1.12) |
| Obesity | **-4.35 (-5.93, -2.77)** |  | 0.38 (-1.16, 1.91) | 0.37 (-1.17, 1.92) | 0.38 (-1.17, 1.93) |
| Excessive alcohol use | **2.91 (1.22, 4.60)** |  | **1.98 (0.49, 3.46)** | **1.98 (0.50, 3.47)** | **1.98 (0.50, 3.47)** |
| Time in bed, h | 0.49 (-0.24, 1.23) |  |  | 0.15 (-0.59, 0.88) | 0.14 (-0.60, 0.87) |
| Sleep efficiency, % | **0.43 (0.29, 0.57)** |  |  | 0.06 (-0.11, 0.22) | 0.05 (-0.11, 0.22) |
| Sleep regularity index, per SD | **2.80 (2.01, 3.59)** |  |  |  | -0.17 (-1.29, 0.95) |
| **Pooled R² (95% CI)** |  | **0.284 (0.233, 0.336)** | **0.297 (0.246, 0.349)** | **0.298 (0.246, 0.350)** | **0.298 (0.246, 0.350)** |
| **Mean BIC** |  | **6352.7** | **6299.2** | **6312.2** | **6318.8** |
| Regression coefficients and variances were combined across 10 completed RAR datasets using Rubin's rules; 95% CIs and two-sided p values were calculated from the pooled estimates. Positive coefficients indicate higher cognitive T-scores. Boldface indicates pooled nominal p < 0.05 and does not denote Benjamini-Hochberg-adjusted significance. For relative amplitude, Benjamini-Hochberg correction was applied across the 8 secondary outcomes separately within each model. RAR, rest-activity rhythm; FQHC, federally qualified health center. | | | | | |
| Model 1 estimates were obtained from separate unadjusted regressions for each predictor, so a common Model 1 R² or BIC is not applicable. Model 2 included intradaily variability, interdaily stability, age, sex, education, and recruitment site type. Model 3 additionally included hearing impairment, hypercholesterolemia, depressive symptoms, average daily activity, diabetes, current smoking, hypertension, obesity, and excessive alcohol use. Model 4 additionally included time in bed and sleep efficiency; Model 5 additionally included the sleep regularity index. | | | | | |
| Reference categories were non-female sex, academic recruitment site, and absence of each binary risk factor. Predictors labeled 'per SD' were standardized (mean = 0, SD = 1). R² and its 95% CI were pooled across the 10 completed datasets using a Fisher z transformation. BIC is the arithmetic mean of the imputation-specific values and is descriptive; it was not combined using Rubin's rules. | | | | | |

**eTable 11. Associations between relative amplitude and NIH Toolbox expressive language T-scores**

| **Predictor** | **Model 1 Unadjusted** | **Model 2 RAR + sociodemographic N = 851** | **Model 3 + dementia risk factors N = 837** | **Model 4 + time in bed + sleep efficiency N = 837** | **Model 5 + sleep regularity N = 837** |
| --- | --- | --- | --- | --- | --- |
| Relative amplitude, per SD | **2.81 (2.04, 3.57)** | 0.78 (-0.17, 1.73) | 0.66 (-0.32, 1.64) | 0.50 (-0.74, 1.74) | 0.72 (-0.74, 2.18) |
| Intradaily variability, per SD | **1.83 (1.06, 2.61)** | 0.59 (-0.21, 1.39) | 0.74 (-0.07, 1.55) | 0.75 (-0.06, 1.57) | 0.81 (-0.03, 1.65) |
| Interdaily stability, per SD | 0.72 (-0.07, 1.51) | 0.15 (-0.81, 1.11) | 0.26 (-0.73, 1.24) | 0.30 (-0.71, 1.31) | 0.38 (-0.67, 1.42) |
| Age, years | 0.03 (-0.06, 0.13) | **-0.13 (-0.21, -0.04)** | **-0.10 (-0.20, -0.00)** | **-0.10 (-0.20, -0.00)** | -0.10 (-0.19, 0.00) |
| Female sex | **-2.18 (-3.80, -0.55)** | -1.24 (-2.71, 0.22) | -1.37 (-2.89, 0.15) | -1.43 (-2.99, 0.12) | -1.45 (-3.01, 0.10) |
| Education, years | **1.87 (1.61, 2.13)** | **1.23 (0.91, 1.55)** | **1.19 (0.87, 1.52)** | **1.19 (0.86, 1.52)** | **1.20 (0.87, 1.53)** |
| Recruitment site: FQHC | **-10.97 (-12.69, -9.25)** | **-6.12 (-8.22, -4.02)** | **-6.22 (-8.45, -3.98)** | **-6.22 (-8.46, -3.97)** | **-6.21 (-8.46, -3.96)** |
| Hearing impairment | -0.81 (-3.09, 1.48) |  | -0.74 (-2.78, 1.31) | -0.75 (-2.79, 1.30) | -0.77 (-2.82, 1.28) |
| Hypercholesterolemia | **-1.78 (-3.38, -0.17)** |  | -0.48 (-2.11, 1.16) | -0.49 (-2.14, 1.15) | -0.47 (-2.11, 1.18) |
| Depressive symptoms | -1.48 (-3.31, 0.35) |  | 1.29 (-0.39, 2.98) | 1.31 (-0.38, 3.00) | 1.26 (-0.44, 2.96) |
| Average daily activity, per SD | -0.10 (-0.89, 0.69) |  | 0.29 (-0.46, 1.04) | 0.35 (-0.46, 1.17) | 0.34 (-0.48, 1.15) |
| Diabetes | **-5.14 (-6.98, -3.29)** |  | **-2.22 (-4.11, -0.32)** | **-2.18 (-4.09, -0.28)** | **-2.17 (-4.07, -0.26)** |
| Smoking | **-6.11 (-8.48, -3.74)** |  | 0.46 (-1.95, 2.88) | 0.51 (-1.92, 2.93) | 0.46 (-1.98, 2.89) |
| Hypertension | **-2.10 (-3.70, -0.50)** |  | 0.78 (-0.80, 2.37) | 0.79 (-0.80, 2.37) | 0.76 (-0.83, 2.35) |
| Obesity | **-3.72 (-5.29, -2.15)** |  | 0.17 (-1.41, 1.74) | 0.16 (-1.42, 1.75) | 0.18 (-1.41, 1.76) |
| Excessive alcohol use | **2.65 (0.98, 4.32)** |  | **1.61 (0.09, 3.13)** | **1.61 (0.08, 3.13)** | **1.61 (0.09, 3.13)** |
| Time in bed, h | 0.46 (-0.27, 1.19) |  |  | 0.08 (-0.67, 0.83) | 0.06 (-0.69, 0.81) |
| Sleep efficiency, % | **0.38 (0.24, 0.52)** |  |  | 0.04 (-0.13, 0.20) | 0.03 (-0.14, 0.20) |
| Sleep regularity index, per SD | **2.34 (1.56, 3.11)** |  |  |  | -0.33 (-1.48, 0.82) |
| **Pooled R² (95% CI)** |  | **0.239 (0.190, 0.289)** | **0.249 (0.200, 0.301)** | **0.250 (0.200, 0.301)** | **0.250 (0.200, 0.301)** |
| **Mean BIC** |  | **6430.2** | **6372.1** | **6385.4** | **6391.8** |
| Regression coefficients and variances were combined across 10 completed RAR datasets using Rubin's rules; 95% CIs and two-sided p values were calculated from the pooled estimates. Positive coefficients indicate higher cognitive T-scores. Boldface indicates pooled nominal p < 0.05 and does not denote Benjamini-Hochberg-adjusted significance. For relative amplitude, Benjamini-Hochberg correction was applied across the 8 secondary outcomes separately within each model. RAR, rest-activity rhythm; FQHC, federally qualified health center. | | | | | |
| Model 1 estimates were obtained from separate unadjusted regressions for each predictor, so a common Model 1 R² or BIC is not applicable. Model 2 included intradaily variability, interdaily stability, age, sex, education, and recruitment site type. Model 3 additionally included hearing impairment, hypercholesterolemia, depressive symptoms, average daily activity, diabetes, current smoking, hypertension, obesity, and excessive alcohol use. Model 4 additionally included time in bed and sleep efficiency; Model 5 additionally included the sleep regularity index. | | | | | |
| Reference categories were non-female sex, academic recruitment site, and absence of each binary risk factor. Predictors labeled 'per SD' were standardized (mean = 0, SD = 1). R² and its 95% CI were pooled across the 10 completed datasets using a Fisher z transformation. BIC is the arithmetic mean of the imputation-specific values and is descriptive; it was not combined using Rubin's rules. | | | | | |

**eTable 12. Multicollinearity diagnostics for Model 4 across cognitive outcomes**

| **Predictor** | **Fluid composite** | **Attention** | **Executive function** | **Episodic memory** | **Working memory** | **Processing speed** | **Crystallized composite** | **Receptive language** | **Expressive language** |
| --- | --- | --- | --- | --- | --- | --- | --- | --- | --- |
| Relative amplitude (z) | 1.77 | 1.79 | 1.78 | 1.78 | 1.79 | 1.79 | 1.78 | 1.78 | 1.79 |
| Intradaily variability (z) | 1.18 | 1.17 | 1.17 | 1.18 | 1.17 | 1.17 | 1.17 | 1.17 | 1.17 |
| Interdaily stability (z) | 1.43 | 1.45 | 1.44 | 1.44 | 1.45 | 1.45 | 1.44 | 1.44 | 1.45 |
| Age, years | 1.14 | 1.13 | 1.13 | 1.13 | 1.14 | 1.13 | 1.13 | 1.13 | 1.13 |
| Female sex | 1.08 | 1.08 | 1.08 | 1.08 | 1.08 | 1.08 | 1.08 | 1.08 | 1.08 |
| Education, years | 1.27 | 1.29 | 1.29 | 1.28 | 1.28 | 1.29 | 1.28 | 1.29 | 1.29 |
| Recruitment site | 1.35 | 1.36 | 1.36 | 1.36 | 1.36 | 1.36 | 1.36 | 1.36 | 1.36 |
| Hearing impairment | 1.02 | 1.02 | 1.02 | 1.02 | 1.02 | 1.02 | 1.02 | 1.02 | 1.02 |
| Hypercholesterolemia | 1.16 | 1.16 | 1.16 | 1.16 | 1.16 | 1.16 | 1.16 | 1.16 | 1.16 |
| Depressive symptoms | 1.04 | 1.05 | 1.05 | 1.05 | 1.05 | 1.05 | 1.05 | 1.05 | 1.05 |
| Average daily activity (z) | 1.16 | 1.17 | 1.17 | 1.16 | 1.17 | 1.17 | 1.17 | 1.17 | 1.17 |
| Diabetes | 1.15 | 1.15 | 1.15 | 1.15 | 1.15 | 1.15 | 1.15 | 1.15 | 1.15 |
| Smoking | 1.13 | 1.14 | 1.14 | 1.14 | 1.14 | 1.14 | 1.14 | 1.14 | 1.14 |
| Hypertension | 1.12 | 1.12 | 1.12 | 1.12 | 1.12 | 1.12 | 1.12 | 1.12 | 1.12 |
| Obesity | 1.13 | 1.13 | 1.13 | 1.13 | 1.13 | 1.13 | 1.13 | 1.13 | 1.13 |
| Excessive alcohol use | 1.03 | 1.03 | 1.03 | 1.03 | 1.03 | 1.03 | 1.03 | 1.03 | 1.03 |
| Time in bed, h | 1.18 | 1.17 | 1.18 | 1.18 | 1.17 | 1.17 | 1.17 | 1.17 | 1.17 |
| Sleep efficiency, % | 1.35 | 1.36 | 1.36 | 1.35 | 1.36 | 1.36 | 1.36 | 1.36 | 1.36 |
| Values are the maximum adjusted generalized variance inflation factors (GVIF^(1/(2 x Df))) across the 10 imputation-specific models. These diagnostic statistics were not pooled using Rubin's rules. For single-Df continuous predictors, adjusted GVIF equals the square root of VIF. A VIF threshold of 5 corresponds to an adjusted GVIF threshold of approximately 2.24. Columns differ slightly in sample size because each outcome has its own fixed complete-case participant set. | | | | | | | | | |
| (z) indicates that the variable was standardized (mean = 0, SD = 1) before model fitting. | | | | | | | | | |

**eTable 13. Multicollinearity diagnostics for Model 5 across cognitive outcomes**

| **Predictor** | **Fluid composite** | **Attention** | **Executive function** | **Episodic memory** | **Working memory** | **Processing speed** | **Crystallized composite** | **Receptive language** | **Expressive language** |
| --- | --- | --- | --- | --- | --- | --- | --- | --- | --- |
| Relative amplitude (z) | 2.06 | 2.11 | 2.08 | 2.09 | 2.11 | 2.11 | 2.08 | 2.08 | 2.11 |
| Intradaily variability (z) | 1.21 | 1.21 | 1.21 | 1.21 | 1.21 | 1.21 | 1.21 | 1.21 | 1.21 |
| Interdaily stability (z) | 1.47 | 1.50 | 1.49 | 1.48 | 1.49 | 1.50 | 1.48 | 1.48 | 1.50 |
| Age, years | 1.14 | 1.14 | 1.14 | 1.14 | 1.14 | 1.14 | 1.14 | 1.14 | 1.14 |
| Female sex | 1.08 | 1.08 | 1.08 | 1.08 | 1.08 | 1.08 | 1.08 | 1.08 | 1.08 |
| Education, years | 1.28 | 1.29 | 1.29 | 1.28 | 1.29 | 1.29 | 1.29 | 1.29 | 1.29 |
| Recruitment site | 1.35 | 1.36 | 1.36 | 1.36 | 1.36 | 1.36 | 1.36 | 1.36 | 1.36 |
| Hearing impairment | 1.02 | 1.02 | 1.02 | 1.02 | 1.02 | 1.02 | 1.02 | 1.02 | 1.02 |
| Hypercholesterolemia | 1.16 | 1.16 | 1.16 | 1.16 | 1.16 | 1.16 | 1.16 | 1.16 | 1.16 |
| Depressive symptoms | 1.05 | 1.05 | 1.05 | 1.05 | 1.05 | 1.05 | 1.05 | 1.05 | 1.05 |
| Average daily activity (z) | 1.16 | 1.17 | 1.17 | 1.16 | 1.17 | 1.17 | 1.17 | 1.17 | 1.17 |
| Diabetes | 1.15 | 1.15 | 1.15 | 1.15 | 1.15 | 1.15 | 1.15 | 1.15 | 1.15 |
| Smoking | 1.13 | 1.15 | 1.14 | 1.14 | 1.14 | 1.15 | 1.15 | 1.15 | 1.15 |
| Hypertension | 1.12 | 1.12 | 1.12 | 1.12 | 1.12 | 1.12 | 1.12 | 1.12 | 1.12 |
| Obesity | 1.13 | 1.13 | 1.13 | 1.13 | 1.13 | 1.13 | 1.13 | 1.13 | 1.13 |
| Excessive alcohol use | 1.03 | 1.03 | 1.03 | 1.03 | 1.03 | 1.03 | 1.03 | 1.03 | 1.03 |
| Time in bed, h | 1.19 | 1.17 | 1.18 | 1.18 | 1.17 | 1.17 | 1.17 | 1.17 | 1.17 |
| Sleep efficiency, % | 1.37 | 1.38 | 1.37 | 1.37 | 1.38 | 1.38 | 1.38 | 1.38 | 1.38 |
| Sleep regularity index (z) | 1.60 | 1.66 | 1.64 | 1.63 | 1.65 | 1.66 | 1.63 | 1.63 | 1.66 |
| Values are the maximum adjusted generalized variance inflation factors (GVIF^(1/(2 x Df))) across the 10 imputation-specific models. These diagnostic statistics were not pooled using Rubin's rules. For single-Df continuous predictors, adjusted GVIF equals the square root of VIF. A VIF threshold of 5 corresponds to an adjusted GVIF threshold of approximately 2.24. Columns differ slightly in sample size because each outcome has its own fixed complete-case participant set. | | | | | | | | | |
| (z) indicates that the variable was standardized (mean = 0, SD = 1) before model fitting. | | | | | | | | | |

**eTable 14. Interactions between relative amplitude and age, sex, and recruitment site type**

| **Moderator** | **Cognitive outcome** | **N** | **Interaction coefficient (95% CI)** | **P value** | **BH-adjusted P value** |
| --- | --- | --- | --- | --- | --- |
| Age | Fluid composite | 817 | -0.01 (-0.09, 0.07) | 0.83 |  |
| Age | Attention | 837 | 0.02 (-0.07, 0.11) | 0.68 | 0.77 |
| Age | Executive function | 836 | 0.02 (-0.05, 0.09) | 0.56 | 0.74 |
| Age | Episodic memory | 825 | -0.06 (-0.15, 0.02) | 0.15 | 0.33 |
| Age | Working memory | 832 | 0.05 (-0.03, 0.12) | 0.21 | 0.33 |
| Age | Processing speed | 837 | 0.01 (-0.07, 0.08) | 0.81 | 0.81 |
| Age | Crystallized composite | 832 | 0.10 (0.00, 0.20) | 0.041 | 0.17 |
| Age | Receptive language | 833 | 0.06 (-0.03, 0.15) | 0.17 | 0.33 |
| Age | Expressive language | 837 | 0.09 (0.00, 0.18) | 0.042 | 0.17 |
| Sex | Fluid composite | 817 | -1.14 (-2.46, 0.18) | 0.09 |  |
| Sex | Attention | 837 | -1.12 (-2.57, 0.34) | 0.13 | 0.29 |
| Sex | Executive function | 836 | -1.09 (-2.24, 0.06) | 0.06 | 0.29 |
| Sex | Episodic memory | 825 | -0.04 (-1.45, 1.38) | 0.96 | 0.96 |
| Sex | Working memory | 832 | -0.28 (-1.51, 0.94) | 0.65 | 0.74 |
| Sex | Processing speed | 837 | -0.59 (-1.84, 0.65) | 0.35 | 0.47 |
| Sex | Crystallized composite | 832 | 1.17 (-0.41, 2.76) | 0.15 | 0.29 |
| Sex | Receptive language | 833 | 1.05 (-0.36, 2.47) | 0.14 | 0.29 |
| Sex | Expressive language | 837 | 0.90 (-0.53, 2.33) | 0.22 | 0.35 |
| Recruitment site | Fluid composite | 817 | -0.94 (-2.41, 0.53) | 0.21 |  |
| Recruitment site | Attention | 837 | 0.18 (-1.44, 1.79) | 0.83 | 0.87 |
| Recruitment site | Executive function | 836 | -0.11 (-1.39, 1.17) | 0.87 | 0.87 |
| Recruitment site | Episodic memory | 825 | -0.53 (-2.10, 1.05) | 0.51 | 0.68 |
| Recruitment site | Working memory | 832 | -1.57 (-2.92, -0.21) | 0.024 | 0.09 |
| Recruitment site | Processing speed | 837 | -0.75 (-2.13, 0.63) | 0.29 | 0.46 |
| Recruitment site | Crystallized composite | 832 | -1.88 (-3.64, -0.12) | 0.036 | 0.10 |
| Recruitment site | Receptive language | 833 | -1.89 (-3.45, -0.33) | 0.018 | 0.09 |
| Recruitment site | Expressive language | 837 | -1.01 (-2.60, 0.58) | 0.21 | 0.43 |
| Each interaction was added separately to Model 4. Interaction coefficients and variances were combined across 10 completed RAR datasets using Rubin's rules; 95% CIs and two-sided p values were calculated from the pooled estimates. | | | | | |
| The Fluid Cognition Composite was the prespecified primary outcome and was not included in the correction family. Benjamini-Hochberg correction was applied to the pooled p values separately for each moderator across the 8 secondary outcomes. | | | | | |
| Reference categories were non-female sex and academic recruitment site. | | | | | |

**eFigure 2. Adjusted marginal mean NIH Toolbox cognitive domain T-scores by relative amplitude quartile**

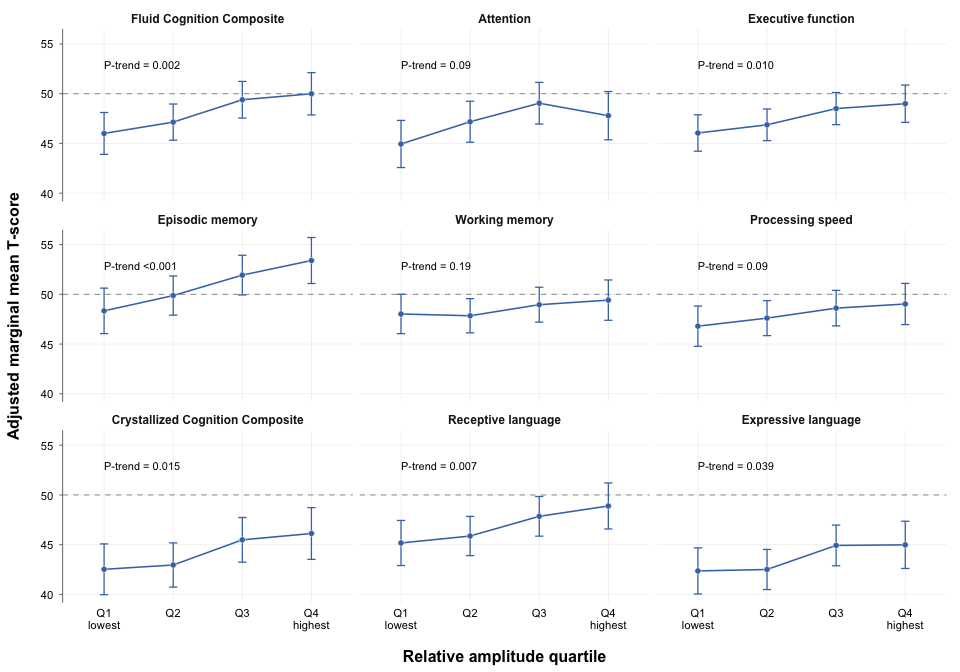

Points and error bars show adjusted marginal means and 95% CIs from fully adjusted Model 4, combined across the 10 completed RAR datasets using Rubin's rules. The dashed horizontal line marks the NIH Toolbox normative T-score mean of 50. Two-sided P values for trend were calculated by modeling relative amplitude quartile as an ordinal variable within each imputation and pooling the coefficient and variance using Rubin's rules. The Benjamini-Hochberg procedure was applied to the pooled trend P values across the eight secondary cognitive outcomes.
